# Combinatorial effects of gene dosage, polygenic background and environment on complex traits

**DOI:** 10.64898/2026.04.30.26352063

**Authors:** Molly F. Sacks, Marieke Klein, Tim B. Bigdeli, Mart Kals, Matthew T. Oetjens, Florian Bénitière, Jacquelyn Johnson, Adam Maihofer, Margit Nõukas, Michael Francis, Bryan Gorman, Iskander Said, Giulio Genovese, Georgios Voloudakis, Kyriacos Markianos, Murray Stein, Joel Gelernter, David H. Ledbetter, Caroline M. Nievergelt, Christa Lese Martin, Vincent-Raphaël Bourque, Omar Shanta, Jeffrey R. MacDonald, Bhooma Thiruvahindrapuram, Mamad Ahangari, Anjali Srinivasan, James Guevara, Jessica H. Hall, Josephine E. Haddon, Claudia Vingerhoets, David Linden, Mieke M. van Haelst, Marianne B.M. van den Bree, Carrie E. Bearden, Raquel E. Gur, T. Blaine Crowley, Daniel E. McGinn, Beverly S. Emanuel, Elaine H. Zackai, Ann Swillen, Thérèse van Amelsvoort, Jacob Vorstman, Anne S. Bassett, Donna M. McDonald-McGinn, Panos Roussos, Mihaela Aslan, Philip D. Harvey, Million Veteran Program, Estonian Biobank Research Team, Genes to Mental Health Network, Sébastien Jacquemont, Saiju Pyarajan, Kelli Lehto, Peter M. Visscher, Jonathan Sebat

## Abstract

Complex traits arise from the combined effects of rare and common genetic variation, development and environment, but resolving their joint contributions has been limited by statistical power. Here, we meta-analyze effects of recurrent copy number variants (CNVs), polygenic scores, sex, age and medications on height and body mass index in 1,447,001 individuals across 6 biobanks and clinical cohorts. CNVs show largely mirror dose-dependent effects of deletions and duplications on both traits, but a subset of loci exhibit asymmetric dose-responses on adult height, consistent with buffering of one allele but not the other. Polygenic background and medications combine with CNVs in ways broadly consistent with additivity. However, detailed analyses of loci at 16p11.2 and 22q11.2 reveal context-dependent effects that vary across development, physiology and sex. At 22q11.2, the net effect of a CNV reflects opposing and reinforcing contributions of multiple genes, providing a potential mechanism for buffering of dosage effects. These results indicate that genetic effects follow additive patterns in aggregate, while context-dependent deviations are widespread for specific loci.

## Main

Complex traits such as height^1–3^ and body mass index (BMI)^1,4,5^ are shaped by many genetic and environmental influences, both rare and common. Yet the principles by which these factors combine within individuals remain poorly understood. Genome-wide association studies (GWAS) have characterized polygenic architecture,^6–9^. Rare variants, such as copy number variants (CNVs)^10–12^, loss-of-function^13–15^ and damaging missense^13^ variants, also have a significant influence. Research has begun to explore the effects of rare and common variants with each other^16–19^ and with the environment^20,21^. The major problem has been the limitation in statistical power. Common variants cover a vast search space with individually modest effects, while rare variants of large effect are found in very small numbers within individual datasets, limiting power to estimate effect sizes and interactions.

Recurrent copy number variants (CNVs) provide one solution to this problem. Generated by non-allelic homologous recombination (NAHR), recurrent deletions (DELs) and duplications (DUPs) repeatedly perturb gene dosage at the same genomic locus in both directions^22^. At some genomic regions, multiple breakpoints generate nested or overlapping CNVs, resulting in dosage perturbations of adjacent loci individually and in combination. This modular architecture enables a systematic dissection of how genes act in combination to influence complex traits. Lastly, the high per-locus mutation rates of recurrent CNVs result in a comparatively high population prevalence of these large-effect variants^23^, and their detectability with SNP-genotyping arrays allows population-scale studies, enabling greater power to detect effects.

This study investigates how recurrent copy number variants (CNVs) influence height and body mass index (BMI) and how their effects vary within the context of polygenic background, sex, development and environment. To this end, we performed a federated meta-analysis of CNV and SNP genotypes, where a harmonized analysis workflow for CNV genotyping, polygenic scoring, phenotype tabulation, scaling, and association testing was applied across five biobanks: UK Biobank (n = 341,425), MyCode (n = 110,497), Million Veteran Program (n = 587,744), Estonian Biobank (n = 63,220), All of Us (n = 344,075), and clinically-ascertained cohorts (N = 9,652), for a total sample of 1,447,001 individuals. (**Fig. 1**).

**Figure 1:**
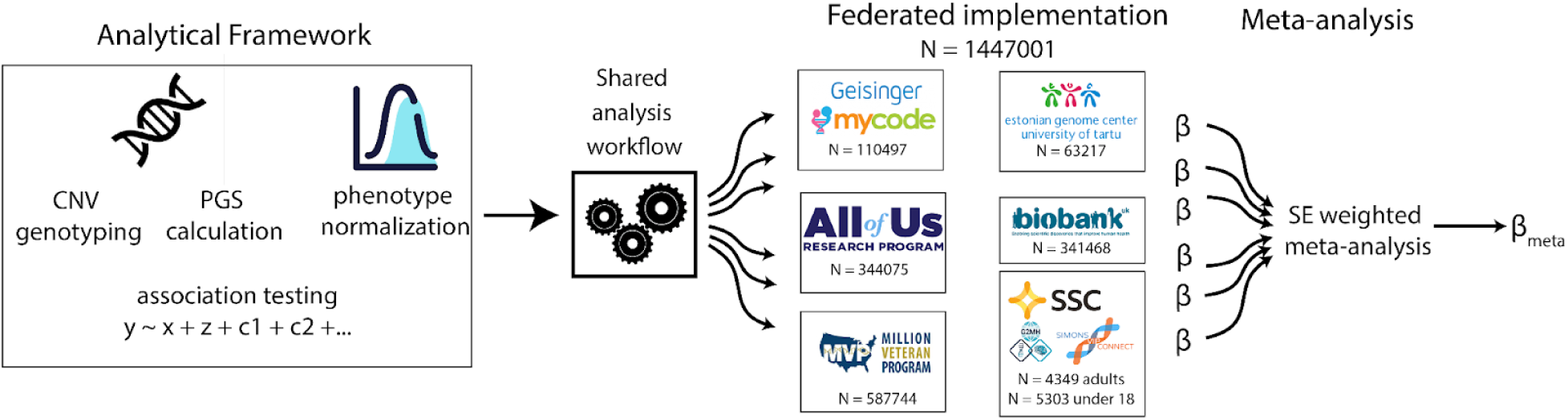
Federated biobank meta-analysis of effects of recurrent CNVs and PGS on height and BMI.

### Rare CNVs have large, dose-dependent effects on BMI and Height

Genome-wide analysis of CNVs^24,25^ typically aggregates signals across genes or regions, collapsing diverse rare alleles into a single test and limiting the ability to resolve the effects of gene dosage. Here we targeted a predefined set of 94 recurrent DEL and DUP alleles at 47 CNV hotspots that are readily detectable by all genotyping platforms (**Supplementary Table 1**), and many of which have associations with a variety of health or developmental conditions^26^. A focused approach provided precise estimation of genetic effects, reduced complexity of analysis workflows, and maximized statistical power (see Supplementary Note 2). We observed 81 CNV alleles at 47 loci in at least 10 subjects in our combined sample and were detected at comparable frequencies in all five biobanks (**Supplementary Table 2**) (excluding CLIN cohort contained participants which were ascertained on CNV carrier status).

Associations with sex- and age-normalized height and BMI were tested for specific CNV alleles (see Online Methods). For height, 19 CNVs had significant effects after correction for 47 tests (23%, 12 DELs and 7 DUPs, 4 loci had effects for both alleles, **Supplementary Table 3**), with many more nominally significant hits. For BMI, 16 CNVs had significant effects after correction for 47 tests (20%, 8 DELs and 8 DUPs, with 5 loci having effects for both alleles, **Supplementary Table 3**). Additionally, 6 CNVs had significant effects on both traits. Effect size estimates were consistent across biobanks and ancestry groups (**Fig 2A-B**). Results show that these CNVs have comparatively large effects relative to single genes that have been implicated by WES, (**Fig 2C-D**), consistent with a perturbation of multiple genes having a larger effect. However, there was not a significant correlation between CNV length or the number of genes with effect size, suggesting that the effect size of a CNV depends on the specific combinations of genes within it (**Extended Data Fig. 8**).

**Figure 2:**
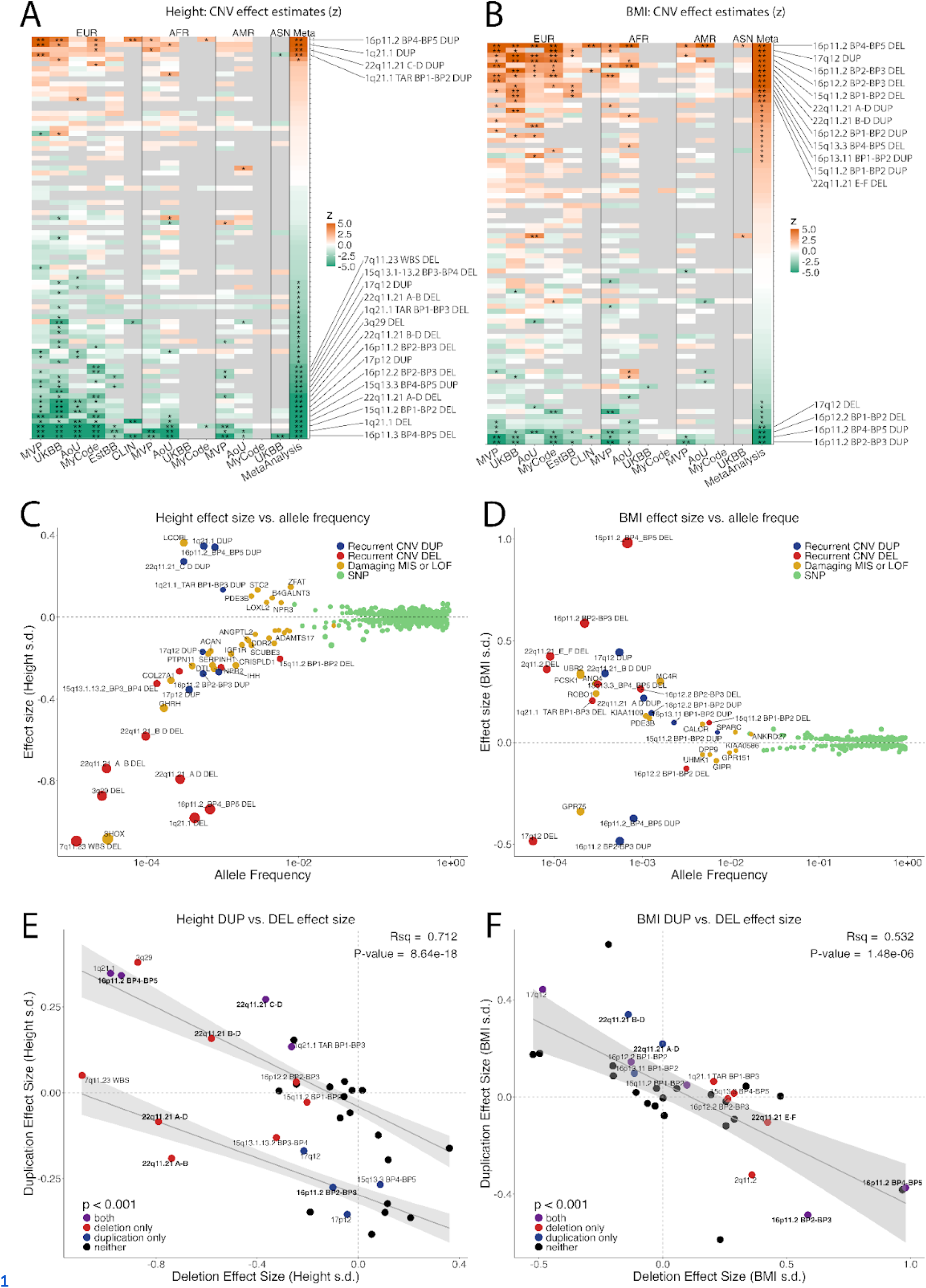
Main effects of recurrent CNVs on body mass index and height. CNV effect sizes (z) for (**A**) height and (**B**) BMI across biobanks and in meta-analysis. **significant after correction for 47 loci, *p < 0.05. Recurrent CNV associations with (**C**) height and (**D**) BMI from the present study (Supplementary Table3) in context with SNPs^1 3^, and rare pLOF or damaging missense variants^1413^. The lead GWS SNP in each 500kb window is shown. Dose-response curves showing correlation of effects sizes of duplication vs. deletion for (**E**) height and (**F**) BMI, which includes 34 loci that had sufficient power (N>10 carriers for both DEL and DUP) to estimate both. Labeled points represent bonferroni significant associations. Summary statistics for panels A-F are in Tables S2 and S3. (E) Recurrent alleles of the 22q11.2 locus are labeled in **bold** to highlight those with mirror effects (22q11.2 C-D and B-D) and asymmetric effects (22q11.2 A-B, A-D) which are described in greater detail in Figure 6.

For both traits, effects of gene copy number were broadly dose-dependent, as evident from a dose-response curve showing a negative correlation of effect sizes for DELs and DUPs across all loci that were powered for both (N >10 carriers for each allele, **Fig. 2E-F**). Comparison of model fits showed that dose responses for height consisted of 2 (linear) components (**Fig. 2E**, **Fig. S1**), one with a “mirror” dose-dependent effect of DEL and DUP and another consisting of loci where effects were negative for one allele and null for the other. This asymmetry suggests that a subset of loci for height are permissive of a negative effect for one allele (DEL or DUP), but are buffered against a positive effect of the other. By contrast, model fits for BMI were consistent with a single linear dose-response curve (**Fig. SZ**).

As expected, recurrent CNVs explain a small fraction of the overall heritability in the population. The CNV loci described here explained 0.23% and 0.18% of the variance in height and BMI respectively (see Methods)(**Supplementary Table 4**). This is less than the heritability explained by the genome-wide burden of rare gene variants, which is estimated at 3.7% and 1.2% for height and BMI^27^, respectively, but it is a non-trivial proportion considering that the CNVs investigated in this study occur in only 3% of our sample and collectively involve just 1.31% (42 Mb) of the genome.

### Combinatorial effects of rare variants, polygenic scores (PGS) and psychiatric medications

Rare and common variants act in combination to influence complex traits such as intellectual disability^28^, autism spectrum disorder^17^, schizophrenia^16,28^, and heart disease^18^. In this study, accounting for the combination of CNV genotype and PGS explained substantial variation. For instance, upon stratifying the 16p11.2 BP4-BP5 locus by PGS quartile, the combined effects explain a range of 19.76 cm for height (**Fig. 3A**) and 14.27 kg/m^2^ for BMI (**Fig. 3B**, **table 5**), compared to a range of 10.87 cm and 6.09 kg/m^2^ (**Table 3**) for the CNV alone.

**Figure 3.**
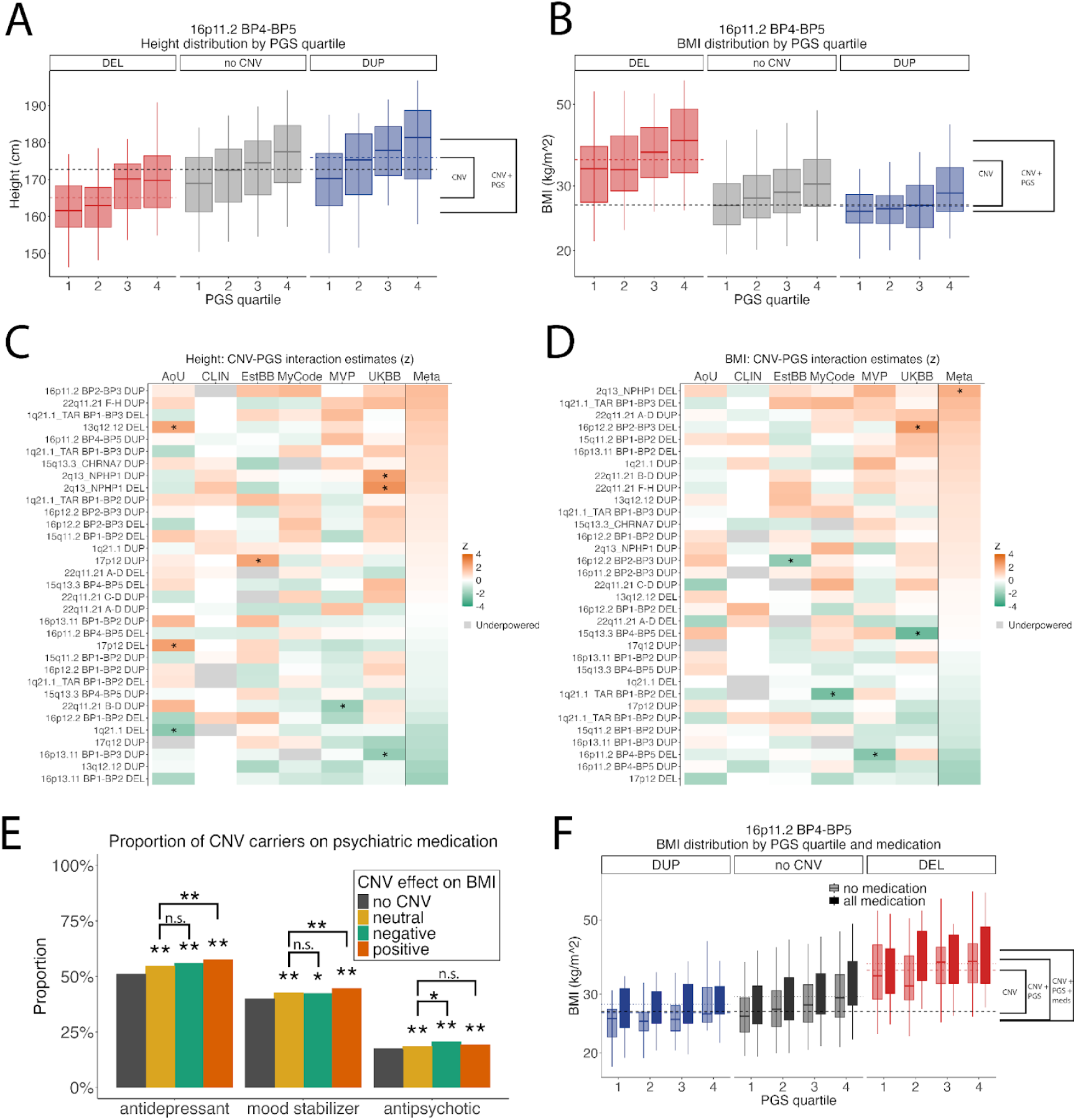
The combined effects of CNV and PGS and medication on complex traits. We also observe wide . variation in traits when we stratified the 16p11.2 locus by CNV genotype and PGS quartile, (A) spanning a range 19.76 cm for height and (B) 14.27 kg/m^2^ for BMI. C-D) Heatmap of z-transformed interaction term estimates in each biobank and the meta-analysis. * = p < 0.05. (E) Proportion of individuals who reported lifetime use of three broad classes of medication, in CNV carriers and non-CNV carriers. ** = p < 0.001 for Chi2. (F) BMI of 16p11.2 BP4-BP5 CNV carriers stratified by PGS quartile and medication. The data used to generate this figure can be found in Supplementary Tables 5 and 6.

Previous studies have characterized the combined effect of rare and common variants. Penetrance and age-at-onset of pathogenic variants in *HNF1A, HNF1B*, and *HNF4A* for type 2 diabetes varied by PGS strata across multiple cohorts^29^. Other studies that have looked at the joint effects of rare and common variants in cancer^30–34^ and cognitive traits^16,17,19,35,36^ confirm that risk within a rare group is higher for the subset with high PGS, but statistically meaningful interactions have not been found.

Our combined data provides unprecedented power to test for non-additive effects of CNVs and PGS (**Supplementary Note 2**). We tested CNV×PGS interactions for the 32 recurrent CNVs in our study that exceeded 200 carriers in the combined cohort. A variety of weak interactions (P<0.05) were observed in single cohorts (8 for height and 5 for BMI), but did not replicate in other cohorts or in the meta-analysis (**Fig. 3C-D**). One nominal association in the meta-analysis (2q13 *NPHP1* DUP × PGS-BMI) was not significant after multiple test correction for 32 tests. We did observe better model fit for models that included interaction terms (**Table 4**); however, the magnitude of these interactions are in general too small to be reliably detected even in a sample of 1.4 million. Thus, the combined effects of CNVs and PGS on complex traits were largely consistent with an additive model.

A number of recurrent CNVs confer susceptibility to adult psychiatric conditions, including major depression, bipolar disorder, and schizophrenia^24^. In turn, many treatments for these disorders, such as antidepressants^37^, mood stabilizers, and antipsychotics^38^ can promote weight gain, potentially exacerbating the effect of a CNV on BMI. We extended this approach to investigate the combined effects of genes, PGS and psychiatric medications (See Online methods for description of how medication data was tabulated, see **Supplementary Table 8** for medication definitions). While CNV effects on BMI were not mediated by medication use (**Extended Data Fig. 3**), these three classes of medications were prescribed more frequently to CNV carriers than to the broader cohort. The prevalence of reported lifetime use of antidepressants and mood stabilizers was greatest for the CNVs that contribute to increased BMI (**Fig. 3E**). However, we did not detect significant gene-environment interactions. The influence of medication on BMI was comparable across CNVs grouped by main effect (Sup **Fig. 3G**) and across quartiles of PGS (Sup **Fig 3H**). Nonetheless, the additive combinatorial effects of CNV, PGS and medications were substantial. Using the example of 16p11.2 BP4-BP5, when stratifying subjects by psychiatric medications, PGS and CNV genotype, their combined effects explained a further 1.227 kg/m^2^ difference in BMI. (**Fig. 3F**).

### Female-biased genetic effects of rare variants on BMI

Previous studies have identified rare and common variants with sex-specific effects, including SNPs that show stronger effects on body fat distribution in females than in males ^39,40^. Sex-stratified analyses of rare variants have also identified female-specific effects^19,41^. For a subset of CNVs with sufficient sample size (>1000 carriers), we have adequate power to detect modest sex-dependent effects (<0.1 SD; **Supplementary Note 2**), but for most CNVs, sex differences can only be assessed in aggregate.

We compared sex-stratified CNV effects on height (**Fig. 4A**) and BMI (**Fig. 4B**). For height, CNV effects were highly correlated between sexes, with a slope not significantly different from 1 (slope = 0.879, p = 0.143), indicating homogeneous effects in males and females (**Fig. 4A**). For BMI, effects were also strongly correlated in males and females but showed a slope significantly less than 1 (slope = 0.707, p = 7.94×10⁻⁸), with many CNVs having stronger positive effects in females. While most individual CNV × sex differences were too small to detect (**Fig. 4C–D**), one (15q11.2 BP1–BP2 DEL) showed a gene × sex interaction on BMI that was significant after Bonferroni correction (**Fig 4D**). Additionally, when collapsed by effect size, CNVs with positive and neutral effects on BMI had significantly stronger positive effects on BMI in females (p = 4.4e-4 for positive CNVs, p = 5.0e-3 for neutral CNVs) (**Supplementary Table 9**). These results are consistent with a modest female-biased effect of rare variants on BMI.

**Figure 4.**
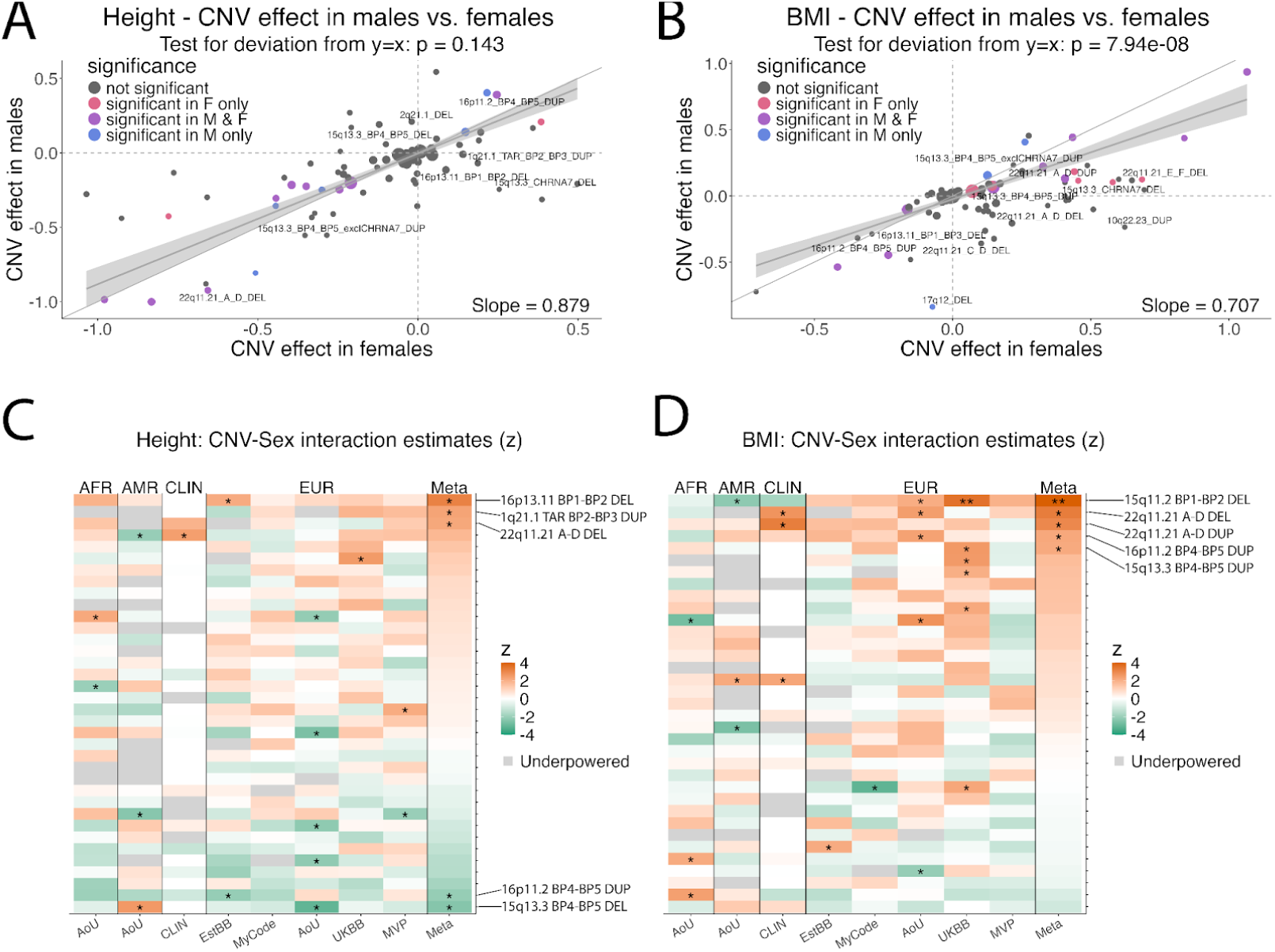
Sex stratified analysis of CNV effects on height and BMI. Correlation of CNV effect sizes in males and females for (**A**) Height and (**B**) BMI. Deviation from y=x was tested via F-test of a joint linear hypothesis. Heatmap of z-transformed interaction term estimates from linear regression in each biobank and the meta-analysis for **C**) Height and (**D**) BMI. Cohorts with significant sex imbalance (such as MVP, 80% male) were excluded. CNVs with under 200 carriers were excluded from interaction analysis (see Supplementary Note 2 on Statistical Power). Summary statistics for panels A-D are in Supplementary Table 9

### Age-dependent effects of CNVs on height are attributable to divergent causal pathways

The results described above represent the effects of recurrent CNVs that are detectable in an adult population. However, phenotypic characterization of CNVs is done almost exclusively in clinically-ascertained pediatric cohorts, which may not be representative of traits in adults. For instance, our results showing a mirror dose-dependent effect of 16p11.2 BP4-BP5 CNVs on height contrast with a previous study in a pediatric sample (ages 5-18) that showed that both the deletion and duplication were associated with short stature^42^.

Using cross-sectional data on participants from our clinically-ascertained cohorts and All of Us, we investigated whether the effects of 16p11.2 CNVs varied across the lifespan. We observed that the effects of the 16p11.2 BP4-BP5 CNVs differed dramatically by age. 16p11.2 BP4-BP5 DUP had divergent effects on height across development consisting of shorter stature as children and taller stature as adults (**Fig. 5A**). The growth trajectory of the reciprocal 16p11.2 BP4–BP5 DEL varied to an even greater extent. The DEL was associated with reduced height early in childhood and again in adulthood, but the trajectory showed transient tall stature between ages 8 and 13. Thus, the conflicting findings of this study and previous reports^42^ are likely explained by age-dependent effects on height (**Fig. 5B**). Likelihood ratio tests comparing nested generalized additive models showed that including a smooth function of height by age significantly improved model fit relative to a null model for both DUP (p=4.51e-5) and DEL (p=5.34e-11) indicating a significant age-dependent effect. Effects of 16p11.2 CNVs on BMI were also age-dependent, but the interaction was weaker (**Extended Data Fig. 4**).

**Figure 5.**
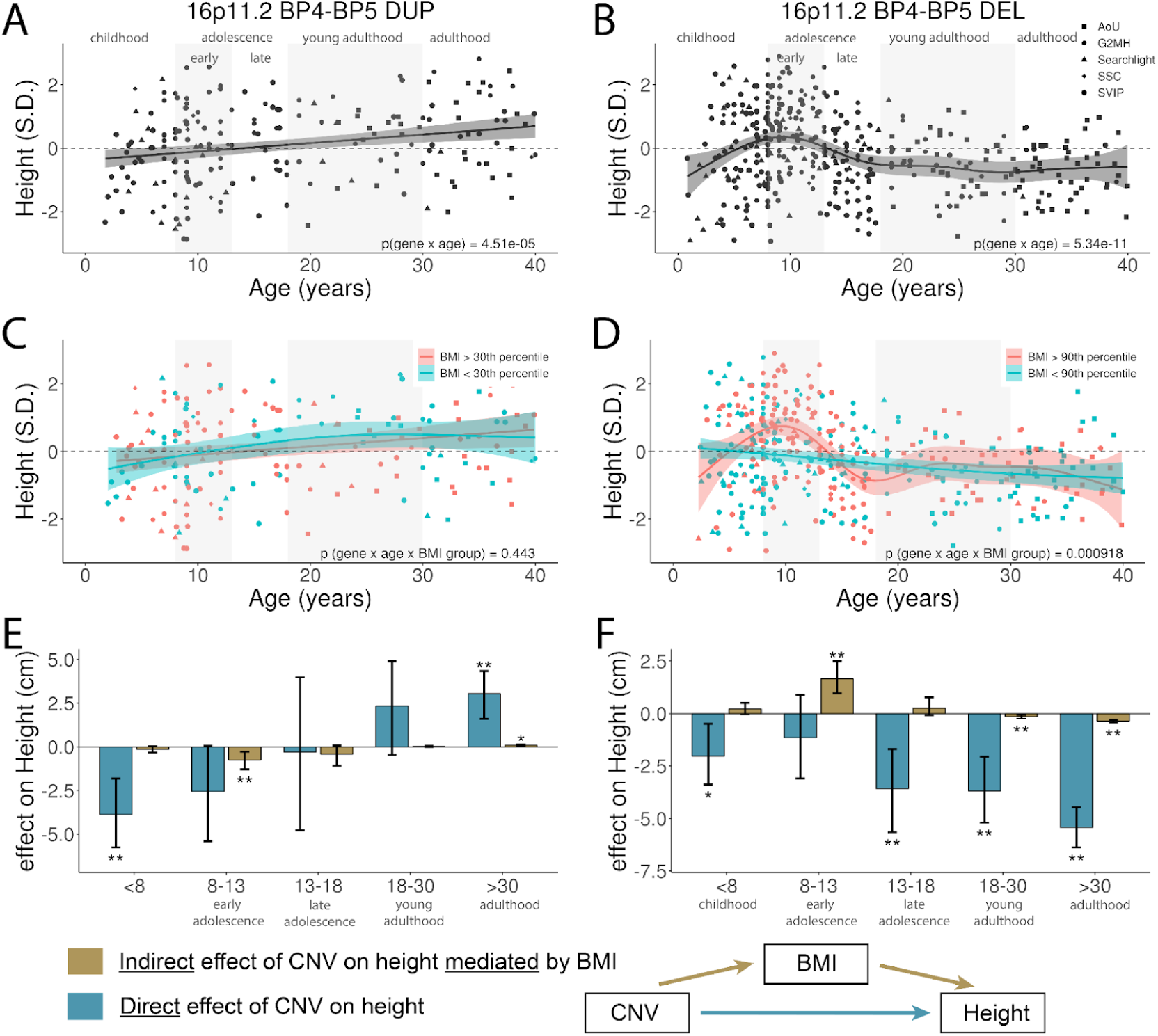
Effects of 16p11.2 BP4-BP5 CNVs differ by age. Age and sex normalized height values vs, age for (A) 16p11.2 BP4-BP5 DUP and (B) 16p11.2 BP4-BP5 DEL. C-D). Cross-sectional height trajectories further stratified by BMI at the 30th (C) and 90th (D) percentiles for DUP and DEL, respectively. E-F). Age-stratified cross-sectional mediation analysis characterizing the distinct causal pathways underlying CNV effects on height, for DUP (E) and DEL (F). Z scores represent height normalized by age and sex relative to WHO norms, and deviation from zero represents the difference in height relative to the normative population. The upper end of each age range is exclusive (i.e., 8-13 represents those who are <u>at least</u> 8 years old but under 13 years old).

Previous research has shown that childhood obesity can influence growth in 16p11.2 deletion syndrome^43^ and in other monogenetic forms of obesity^44^. We investigated two hypotheses to explain age-dependent effects of 16p11.2 BP4–BP5 CNVs on height: first that CNVs have direct effects that vary by age, or second that age-dependencies are attributable to indirect effects on growth mediated by childhood obesity. CNV carriers were split evenly into lean and obese groups by a cutoff at the 90th percentile of BMI (based on population norms). When there were very few obese subjects (for example, the 16p11.2 BP4-BP5 DUP), a less stringent threshold was used to more evenly split the population. For 16p11.2 BP4–BP5 DUP (**Fig. 5C**), age-dependent effects on height did not differ by BMI group (p = 0.451). For the DEL (**Fig. 5D**), the steep growth trajectory was entirely restricted to subjects with obesity (BMI >90th percentile) with a height–age relationship that was significantly different than for subjects without obesity (p = 9.18×10⁻⁴). This differential height-age relationship between subjects with obesity and subjects without obesity was present in both female and male 16p11.2 BP4-BP5 DEL carriers in sex stratified analyses (**Extended Data Fig. 5**). No significant differences were observed when stratified by PGSs (**Extended Data Fig. 5**). The absence of age-dependent effects for PGS-BMI and PGS-Height further supports that these patterns reflect true CNV × age and CNV × BMI × age interactions, rather than artifacts of population stratification in cross-sectional data.

We then performed cross-sectional mediation analysis across developmental stages to differentiate direct CNV effects (CNV → height) from indirect effects (CNV → BMI → height). Indirect effects were observed for both DUP (**Fig. 5E**) and DEL (**Fig. 5F**) during early adolescence. Testing the reverse pathway (CNV → height → BMI) showed no indirect effects for DEL, supporting a true BMI-mediated effect. For DUP, nominal reverse effects suggested the observed mediation may reflect correlation between height and BMI rather than causation. Together, these findings suggest that the DUP has age-dependent effects on height that are mostly a direct effect, and age-dependent effects of the DEL consist with a combination of negative direct effects on height and transient positive effects mediated by childhood obesity.

Age-dependent effects of CNVs on height were then replicated in a clinically-ascertained sample of 22q11.2 A-D DUP (N = 210) and 22q11.2 A-D DEL (N=531) (**Extended Data Fig. 6**). In contrast to its association with short stature in adults (**Fig. 1**), the 22q11.2 DUP was associated with tall stature in children (**Fig. 5A**, p = 0.00012). Likewise, associations of the DEL with short stature varied by age with a stronger effect in children (**Fig. 5B**, p = 0.0258). The DEL effect on Height also showed a significant interaction with BMI (**Fig. 5D**). Our results demonstrate that, for both loci 16p11.2 and 22q11.2, age-dependent effects were attributable to similar mechanisms. Duplications exhibit divergent direct effects on stature in children and adults, while deletions are subject to opposing effects of multiple causal pathways including (1) direct (negative) effects of the deletion and (2) transient (positive) effects mediated by childhood obesity.

### The influence of a CNV represents multiple genes with opposing or reinforcing effects

We further investigated the basis of asymmetric dose responses on height for the 22q11.2 A-D locus (**Fig. 1E**). Its modular genomic architecture enables us to dissect the combinatorial effects of three subregions, each separated by a dense cluster of segmental duplications (**Fig 2A**). NAHR rearrangements can occur between any combination of breakpoints A, B, C and D ^45^, thereby producing DUPs or DELs of the A-B (28 genes), B-C (5 genes) and C-D (12 genes) subregions or multiple adjacent loci (A-C, B-D, A-D) ^46^. By comparing gene dosage effects of each locus individually and together, we can quantify their combinatorial effects.

Previous studies in small cohorts and case reports have reported that 22q11.2 DUPs exhibit both short stature and overgrowth ^47–49^. Here, we show that DUPs of each of the three subregions have distinct effects on height: A–B DUPs were associated with reduced stature, B–C DUPs had no effect, and C–D DUPs were associated with increased stature (**Fig. 6B**). When the neutral B–C interval was co-duplicated with either A–B (**Fig. 6C**) or C–D (**Fig. 6D**), the combined effect was neutral (similar to the largest individual effect). For DUPs spanning all three segments (A–D), the opposing effects of A–B and C–D offset each other, resulting in an attenuated net effect of the A-D DUP on height (**Fig. 6E-F**).

**Figure 6:**
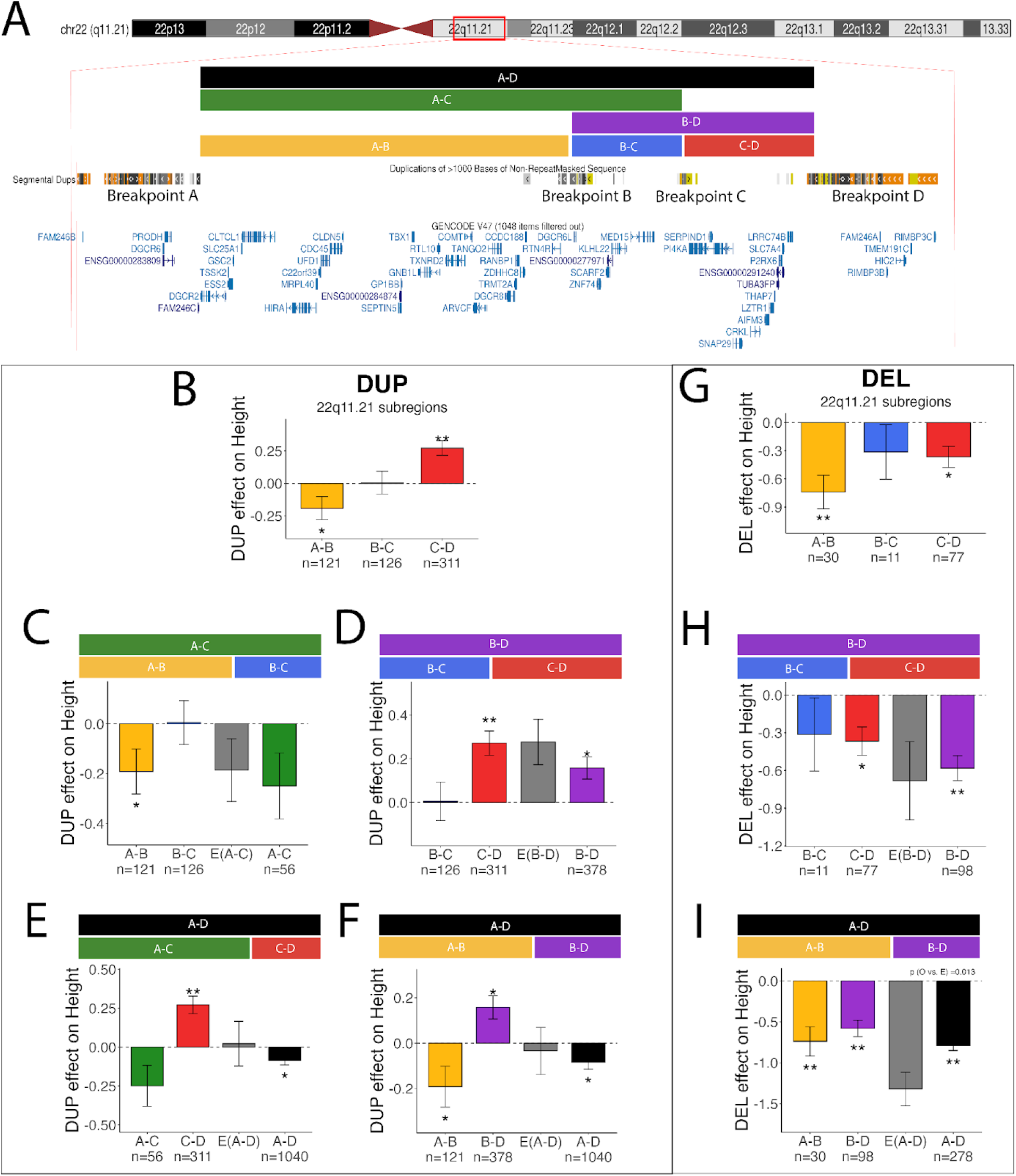
Dissection of the 22q11.2 A-D locus and its effects on height. A). Schematic of the genes encompassed by the most commonly observed CNV breakpoints in our study, as defined by Gencode V49 accessed through the UCSC Genome Browser^56^ B-C). Breakdown of the effects of DUPs (B-F) and DELs (G-I) of 22q11.2 subregions. E(A-C) denotes the expected value of the effect size of A-C based on the sum of the effects of subregions A-B and B-C. Summary statistics for panels B-I are in Table S3.

The effects of 22q11.2 DELs on height have been well characterized previously^46,50–55^. Consistent with these studies, DELs were associated with reduced stature (**Fig. 6G**), and we show that the combined effect of A–B and B–C DELs (A–C) was consistent with an additive model (**Fig. 6G**). However, the effect of deletions spanning the full region (A–D) was weaker than expected under additivity (Observed vs. Expected, P = 0.01; **Fig. 6H**). While the mechanism underlying this sub-additive effect is unclear, it parallels the buffering observed for duplications, suggesting that partially opposing effects among genes within the locus may also attenuate the net impact of deletions on growth.

Analysis of BMI at the same locus reveals a complementary pattern. For DUPs, subregions largely showed convergent, reinforcing effects consistent with additivity (**Extended Data Fig. 7**). In contrast, DELs of the A–B and C–D subregions had opposing effects on BMI (**Extended Data Fig. 7C**), and when combined in the full A–D DEL, these effects partially neutralized one another (**Extended Data Fig. 7I**). Together with the opposing effects observed for duplications on height, these results suggest a buffering of the phenotypic effects of a CNV can arise due to a combination of gene effects with opposing directionalities. These results highlight a potential mechanism by which gene dosage effects are constrained.

## Discussion

A sample of this scale—1.4 million individuals—provides the statistical power required to examine how rare and common variants, together with developmental and environmental factors, shape complex traits. By leveraging recurrent CNVs as modular perturbations of gene dosage, we identify general principles governing their effects. Gene dosage shows largely mirror, dose-dependent relationships with height and BMI, and effects of these CNVs combine additively with polygenic background and medication use. However, analyses within specific contexts reveal a more complex biology, including asymmetric dose responses consistent with buffering, as well as sex-dependent effects on BMI, age-dependent effects on growth, and modulation by physiological state such as obesity.

Recurrent CNVs represent among the largest-effect variants identified in genome-wide studies. The median un-signed effect size for recurrent CNVs was 0.341 SD (2.56 cm for males) compared to 0.177 SD (1.328 cm) for rare gene variants and 0.023 SD (0.172 cm) for common SNPs^57^. Our results are consistent with large-scale copy number changes of multiple genes having large combined effects. However, no clear relationship was found between the number of genes within a locus and its effect size (**Extended Data Fig. 8**); thus, the net effect of a CNV is dependent on the particular combination of genes within it.

While most loci follow a linear “mirror” dose-response with DEL and DUP typically having effects in opposite directions, a subset of loci for height have effects that are asymmetric and biased in the negative direction (short stature), suggesting that the effects of some gene clusters are tightly buffered against positive effects on growth. This result is consistent with another preprint by Milind et al. ^58^, which found evidence that non-monotonic (U-shaped) dose-response curves are common for height, such that both DELs and DUPs typically shift the trait in the negative direction.

Several mechanisms could underlie asymmetric gene-dosage effects, including negative feedback regulation, dosage compensation, and compensatory interactions within gene regulatory networks ^59^. A model proposed by Milind et al.^58^ hypothesized that buffering is intrinsic to the trait, due to the manner in which the trait responds to diverse forms of upstream dysregulation ^58^. This theory likely could explain non-monotonic effects of CNVs on traits such as cognitive performance where reciprocal DELs and DUPs at the same locus are both associated with developmental delay ^60^. However, results in this study support a different mechanism for height (see **Supplementary Note 3** for discussion of results in context of Milind et al.). In our data, only a subset (∼30%) of loci have asymmetric effects, and only a fraction of these can be described as non-monotonic (e.g. 22q11.2 A–B and A–D, **Fig. 6**). We show that buffering can arise from the aggregate behavior of multiple genes within a locus. When opposing effects of the 22q11.2 A-B and C-D DUPs are combined within the full A-D DUP, there is an additive cancelation of opposing effects (**Fig. 6F**). When reinforcing negative effects of the A-B and C-D DELs are combined with the full A-D DEL, the net effect is sub-additive (**Fig. 6I**), suggesting that gene dosage effects can also be attenuated in the negative direction. Together, our findings suggest that buffering can operate in both directions through combinations of reinforcing and opposing gene effects. Because many CNVs in the population span multiple adjacent genes, it is likely that some non-monotonic effects of specific loci described by Milind et al. involve the combinatorial effects of multiple genes.

CNV effects on height were strongly age-dependent, with opposing directional effects in childhood and adulthood. Growth trajectories inferred from cross-sectional data revealed consistent patterns across reciprocal CNVs at both 16p11.2 and 22q11.2. DUPs at both loci showed direct age-dependent effects of genes. At 22q11.2, opposing effects of the A-B and C-D subregions suggest a possible mechanism for age-dependent effects if the A-D DUP: if the effects of genes within the A-B and C-D intervals act at different stages of development, this could produce a positive effect of the A-D DUP in childhood and a negative effect in adulthood. Age-dependent effects of DELs at both loci appear to reflect a combination of causal mechanisms, including negative direct effects on growth and transient positive effects mediated by childhood obesity ^61,44^. This pattern—early overgrowth followed by a plateau—resembles precocious puberty ^62^, a phenotype also associated with 16p11.2 deletion syndrome ^63^. Together, our findings support a model in which age-dependent CNV effects arise from the interplay of direct gene dosage effects on growth with indirect effects of the CNV mediated by metabolic and hormonal pathways.

We observe modest but consistent sex differences in CNV effects on BMI, with stronger positive effects in females, consistent with findings reported in a subset of this sample ^19^. By contrast, a consensus from GWAS has found that SNP effects on height and BMI are generally homogeneous between the sexes^39,64^. Population-level studies of whole genomes or exomes on a similar scale are needed to determine if sex differences are a general property of rare variants of large effect.

While the observed additive effects of polygenic background and medication were not surprising, they were nevertheless informative about the predictability and biological basis of these complex traits. When CNV carriers were stratified by polygenic scores (PGS) the combined effect sizes of the 16p11.2 BP4-BP5 CNV and PGS spanned a range of 19.76 cm for height and 14.27 kg/m^2^ for BMI. These results illustrate how polygenic background shifts the baseline on which large-effect variants act, substantially expanding the range of phenotypic outcomes among carriers.

Medications further shape these effects. Consistent with the associations of recurrent CNVs with psychiatric conditions^24^, CNV carriers were more likely to be prescribed psychiatric medications, particularly those who carried CNVs that predispose to obesity, which is consistent with genetic correlations of BMI and mental health traits^65^. Given that weight gain is a side effect of many antidepressants^37^, mood stabilizers^38^ and antipsychotics^66^, our results provide an example of a pharmacogenomic effect, in which genetic susceptibility to a psychiatric condition itself influences metabolic outcomes of treatment. As we show here, the combined influences of 16p11.2 CNV genotype, PGS and psychiatric medication on BMI are quite large. The stratified group-means span a range of 15.5 kg/m^2^ (**Fig. 3F**), which corresponds to a range of >45 Kg for the average individual.

While this study was designed to define basic principles governing the combinatorial effects of genes in humans, it also has significant clinical implications. Many CNVs studied here are routinely reported in clinical genetic testing (**Supplementary Table 1**) and are associated with developmental, mental health and metabolic conditions^19^. A broad understanding of how CNVs, polygenic background, sex, development and medications jointly shape phenotypic variation can improve the prediction of clinical outcomes and inform patient management. These insights also highlight potential avenues for therapeutic intervention of genetic conditions by identifying modifiable pathways^67^ through which gene dosage can influence development.

## Data Availability

All summary statistics produced in the present study are available upon reasonable request to the authors

## Acknowledgements

This work was funded by grants to J.S. (U01MH119746, OT2OD040415), Department of Veterans Affairs Office of Research and Development, (#I01CX001849), M.T.O (U01MH119705) C.E.B. (R37/R01MH085953, U01MH119736, U01MH124639, R01MH129858, R21MH116473, SFI-AN-AR-HUMAN-00004264-04), J.V. (SFI-AN-AR-HUMAN-00004264-07, R01-MH134965-01; U01-MH119737-01, UO1-MH191719 R01MH106595, BLR&D 1I01BX005920), M.N (Estonian Research Council grant PRG555), A.S. (1U01MH119759), M. B. M. vdB (U01MH119758, U01MH101724, MR/T033045/1, MR/N022572/1, MR/L011166/1, Wellcome Trust Institutional Strategic Support Fund award (503147), The Waterloo Foundation (918-1234), the Baily Thomas Charitable Fund (2315/1), Health & Care Research Wales (Welsh Government, 507556)), P.M.V. (European Research Council #101198904), M.K. (Dutch Research Council (#45219212, #09150162010073), R.E.G. (U01 MH119738; R01MH134969), The Estonian Biobank Research Team (Estonian Center of Genomics/Roadmap II funded by the Estonian Research Council, (project number TT17), K.L. (Estonian Research Council #PSG615, the Ministry of Education and Research Centres of Excellence grant TK218, Estonian Center of Excellence of Well-Being Sciences), P.R. (R01MH125246, R01AG067025, U01MH116442, U24AG087563, VA Department Merit Grant BX004189), C.L.M. and D.H.L. (U01MH119705), S.J. (NI_U01MH119739, CIHR_495906, J-Louis Levesque Chair)

## G2MH

This work was conducted in collaboration with the Genes to Mental Health (G2MH) network, and we acknowledge the contributions of participating investigators and research teams. The G2MH data is available through the NIMH data archive. https://nda.nih.gov/edit_collection.html?id=3228

## All of Us

We gratefully acknowledge *All of Us* participants for their contributions, without whom this research would not have been possible. We also thank the National Institutes of Health’s *All of Us* Research Program for making available the participant data examined in this study.”

## UKBB

This research has been conducted using the *UK Biobank* Resource under application number 62713

## MVP

This research was supported by the Department of Veterans Affairs Cooperative Studies Program (CSP) #572 and the Million Veteran Program (MVP-000,MVP-006 and MVP-076). The MVP is supported by the Office of Research and Development, Department of Veterans Affairs. We thank the MVP staff, researchers and volunteers who have contributed to MVP, and especially those who previously served their country in the military and agreed to enroll in the study (see mvp.va.gov for more information). The contents do not represent the views of the US Department of Veterans Affairs or the US Government. This study was supported by the Veterans Affairs Merit grant BX004189 (to P.R.).

## EstBB

We thank the Estonian Biobank Research Team: Andres Metspalu, Lili Milani, Tõnu Esko, Reedik Mägi, Mait Metspalu,Maris Nelis and Georgi Hudjashov for genotype and health records data.

Grants to Kelli Lehto. *This work was supported by the Estonian Research Council grant PSG615. This work was supported by the Ministry of Education and Research Centres of Excellence grant TK218 Estonian Center of Excellence of Well-Being Sciences*.

The research was conducted using the Estonian Center of Genomics/Roadmap II funded by the Estonian Research Council (project number TT17).

The EstBB data analyses were partially carried out in the High Performance Computing Center, University of Tartu.

EstBB disclaimer under ethics section (or Material & Methods):*The activities of the EstBB are regulated by the Human Genes Research Act, which was adopted in 2000 specifically for the operations of the EstBB. Individual level data analysis in the EstBB was carried out under ethical approval (1.1-12/624 and all further amendments) from the Estonian Committee on Bioethics and Human Research (Estonian Ministry of Social Affairs)*.

## MyCode

This research was conducted using data from the Geisinger MyCode Community Health Initiative and reflects the contributions of participating patients and the Geisinger research team.

## Searchlight (formerly known as Simons VIP)

We are grateful to all of the families at the participating Simons Searchlight sites as well as the Simons Searchlight Consortium, formerly the Simons VIP Consortium.

We appreciate obtaining access to genetic and phenotypic data on SFARI Base.

Approved researchers can obtain the Simons Searchlight population dataset described in this study (v13) by applying at https://base.sfari.org.

## SSC

We are grateful to all of the families at the participating Simons Simplex Collection (SSC) sites, as well as the principal investigators (A. Beaudet, R. Bernier, J. Constantino, E. Cook, E. Fombonne, D. Geschwind, R. Goin-Kochel, E. Hanson, D. Grice, A. Klin, D. Ledbetter, C. Lord, C. Martin, D. Martin, R. Maxim, J. Miles, O. Ousley, K. Pelphrey, B. Peterson, J. Piggot, C. Saulnier, M. State, W. Stone, J. Sutcliffe, C. Walsh, Z. Warren, E. Wijsman). We appreciate obtaining access to phenotypic and genetic data on SFARI Base. Approved researchers can obtain the SSC population dataset described in this study (v15) by applying at https://base.sfari.org.

## Methods

### Genotyping and CNV calling

In previous large-scale studies of CNVs, we have shown that CNV detection varies significantly due to genotyping platform, but confounding due to platform-specific CNV detection is well-controlled when we apply a meta-analytic approach ^24^: (1) all data processing and statistical association tests are first performed within platform, then (2) multiple platforms are combined by meta-analysis of summary statistics. Furthermore, large recurrent CNVs are reliably detected by virtually all commonly-used SNP genotyping platforms using relevant CNV calling software ^24^, and we confirmed that recurrent CNVs were detected in this study with comparable frequencies across all cohorts (**Supplementary Table 2**). Thus, a federated meta-analysis of biobanks can utilize previously existing CNV call sets as input files to standardized shared analysis workflow that is developed centrally and then implemented at all sites. A key aspect of the CNV calling that must be standardized across sites is the assignment of a CNV genotype from raw CNV calls. Methods for standardized CNV genotype assignment is described here and methods for SNP genotyping and raw CNV calling in each Biobank is described below.

#### CNV genotype assignment from existing CNV calls

Each biobank used the same pipeline to generate recurrent CNV genotypes (see Code Availability). To generate recurrent CNV genotypes, we ran bedtools ^68^ intersect, with the CNV calls generated by the various CNV calling methods and the recurrent CNV definitions^24^ (**Supplementary Table 1**) as input. For CNVs with a single set of breakpoints, we require that the observed CNV overlaps with at least 50% of the target CNV region. This corresponds to the -f .5 option in bedtools intersect. For CNV with multiple sets of breakpoints (such as the 22q11.2 region), genotypes were defined as the union of individual sets of breakpoints. For example, for an individual to have a DEL in 22q11.2 A-D region, they would have to have a CNV that spans at least 50% of the A-B, B-C, and C-D regions, and no CNV spanning the D-E, E-F, F-G, or G-H regions.

#### UKBB

Genetic data for the UK Biobank was genotyped on the Affymetrix UK Biobank Axiom Array. CNV calling on Affy Axiom arrays use PennCNV^69^ and QuantiSNP^70^. The consensus of CNV calls from multiple callers was created by merging CNVs at the sample level and retaining CNVs that were called by at least 2 methods. Sample-level and CNV-level QC was performed as previously described^24^.

#### All of Us

A total of 447,278 microarray-derived VCF files were obtained from the All of Us Research Program (https://doi.org/10.1038/s41586-023-06957-x). CNVs were identified using two algorithms: PennCNV^69^ and QuantiSNP^70^. Three samples were excluded due to missing sex information, which is required by both algorithms. Each CNV call set was pre-filtered independently to remove low-confidence variants prior to intersecting the results from both methods. Specifically, CNVs with confidence scores below 15 in either PennCNV or QuantiSNP were excluded. In addition, only CNVs larger than 1 kilobase were retained.

#### EstBB

SNP genotyping data from 205,841 Estonian Biobank (EstBB) participants were generated using the Infinium Global Screening Array (GSA; Illumina Inc.). Samples were excluded if genotype call rate was <95% or if sex inferred from X-chromosome heterozygosity was inconsistent with recorded phenotypic sex. Prior to CNV calling, genotype clusters were manually realigned to improve signal quality, and log R ratio and B allele frequency values were exported from GenomeStudio v2.0.5 (Illumina). Autosomal CNVs were called with cnvPartition v3.2.0 (Illumina Inc.), followed by sample- and CNV-level quality control. Samples with extreme CNV burden (>200 CNV calls or cumulative CNV length >10 Mb) were removed.

#### MyCode

CNVs were called from whole-exome sequencing data using CLAMMS (Copy number estimation using Lattice-Aligned Mixture Models), as described in Maxwell et al. ^71^ and used in subsequent MyCode CNV analyses. SNP genotypes were available for a subset of participants generated on the Illumina HumanOmniExpressExome-8 v1.2 array ^72^. A harmonized set of SNP genotypes on both platform was created using the subset of overlapping variant sites from DiscovEHR WES and the Omni chip platform.

#### MVP

SNP genotyping data for MVP participants was generated using the MVP 1.0 custom Axiom array^73^. Samples were excluded for genotype call missingness >2.5%, excess heterozygosity, potential duplication, or discordance between genetic sex and self-identified gender. Variants were excluded if missingness exceeded 5% or if minor allele frequency differed by more than 10% from 1000 Genomes Project Phase 3 reference data. All analyses were performed using the GRCh38/hg38 human genome reference build. Copy number variants (CNVs) were identified using PennCNV (v1.0.5) with the PennCNV-Affy protocol^74,75^. Raw probe intensity data were converted to log R ratio (LRR) and B-allele frequency (BAF) values using Affymetrix Power Tools, and CNVs were inferred with a hidden Markov model implemented in PennCNV.

#### G2MH

Per-sample data quality was assessed by computing the median absolute deviation (MAD) of GC-corrected coverage (WGS) or Log R Ratio (GSA) values across mappable regions (after excluding assembly gaps, segmental duplications, and simple repeats). Samples with elevated MAD were excluded. For WGS samples, CNVs were called using the DRAGEN CNV caller (v3.8.4, Illumina) and all were manually verified in WGS coverage data. For samples that were genotyped with GSA array samples, CNVs were called with iPattern and PennCNV as described previously ^24^.

#### SSC

Genetic data for the SSC was genotyped on the Illumina 1Mv1, 1Mv3 and HumanOmni2.5-4v1_B microarrays. IDAT files are used to store BeadArray data and Illumina Genome Studio was used to generate genotype call (GTC) files and final report files. CNVs were called with iPattern and PennCNV, and sample-level and CNV-level QC was performed as described previously ^24^.

#### Simons Searchlight

Simons Searchlight (formerly known as Simons VIP ^60^) participants were clinically ascertained on CNV carrier status. Thus, we used the recurrent CNV genotypes provided by SFARI in this study.

### SNP Imputation and QC

SNP Imputation and QC followed slightly different procedures across biobanks, due to each biobank’s unique population structure and genetic data availability. Since individual level data was not combined across biobanks, and ancestry PCs and PGSs were scaled within each biobank, batch effects were not a concern.

For UK Biobank, SNPs were imputed from microarray data using the Axiom array and 1000 Genomes Phase 3 Reference panel. For MyCode, SNPs were imputed from Illumina WES data using the 1000 Genomes Phase 3 Reference panel. For All of Us, SNPs were imputed from Illumina Infinium H3Array data using the Haplotype Reference Consortium reference panel. For EstBB, Imputation was performed with Beagle v5.4 software and default settings. A population-specific reference panel consisting of 2,695 high-coverage (∼30x) WGS samples was utilised for imputation^76^, and standard Beagle hg38 recombination maps were used. MVP was genotyped on MVP 1.0 custom Axiom array, and imputed using the 1000 Genomes Phase 3 Reference panel. For the Clinically Ascertained cohorts, subjects who were only genotyped with SNP microarrays (Infinium GSA-24 for G2MH, HumanOmni2.5-4v1B and Illum1M for SSC, and Affy CytoScan HD and Agilent aCGH for Searchlight) were imputed via the Ricopili^77^ pipeline, with TopMed as the reference panel. SNPs from subjects with WGS data available were then subsetted to match available imputed SNPs. Subjects with no available SNP genotype data (CNV genotypes, demographics, and phenotypes only) were only included in trajectory analyses.

#### Ancestry PCA

Ancestry principal component analysis (PCA) was run separately for each biobank, within broad genetically partitioned ancestry groups (i.e, EUR, AMR, AFR, EAS). We used the flashpca^78^ tool to generate the top 10 ancestry principal components based on a set of minimally correlated SNPs, with exact numbers varying across cohorts. The top 10 PCs explained 1.8% and .25% of variance in height and BMI on average (respectively), though there was mild heterogeneity across cohorts and ancestry groups, with variance explained ranging from .18% to 13% for height and 0.106% to 1.87% for BMI. Across both traits, the AMR ancestry designation had the highest variance explained by PCs. This was true for the AMR ancestry groups in both All of Us and MVP. This is consistent with the fact that this population has a high admixture of haplotypes from broad global ancestry groups. (**Supplementary Table 4**).

#### Polygenic risk score calculation

We used PGScs ^79^ with 25K MCMC iterations, 10K burn-in and phi=0.01 to generate SNP weights based on GWAS: BMI with^4^ and without^1^ UKBB, and height with^2^ and without^3^ UKBB. For downstream analysis, we used the PGSs excluding UKBB for UKBB, and the PGSs including UKBB for all other cohorts. PGSs calculated using PLINK 1.9’s –score option. For downstream analysis, PGSs were scaled to a standard normal distribution for each cohort.

#### Scaling of the response variable

For our adult subjects, we scaled our BMI and Height response variables in a manner consistent with ^5^. We scaled height and BMI in each cohort separately. To scale a response variable y, we fit a model lm(y ∼ sex + age + age^2), and extracted the residuals. Next, we split males and females, then applied a BoxCox transformation to achieve a normal distribution in the response variable. Finally, we recombined males and females. The BoxCox transformation allowed us to achieve normally distributed residuals when fitting out models without moving outliers closer to the mean. This ensures that CNVs with large effect sizes don’t have deflated effect size estimates. Splitting males and females when applying the BoxCox transformation and standard normal scaling ensures that the variance in the response variable is consistent across males and females. A normally distributed response variable is especially important when characterizing interactions, as a non-normal response can result in a misspecified model and lead to type I errors (see Supplementary Note 1: Model Misspecification)

For pediatric subjects, we calculated age and sex norms using the childsds R package^80^, using WHO norms for height and CDC norms for BMI.

#### CNV main effect calculation

To calculate the effect size of each recurrent CNV on height and body mass index, we fit a linear regression model containing CNV genotype coded as a factor (DEL, DUP, and no CNV as the reference) and the top 10 ancestry principal components. For example, to test the effect of CNV A on BMI in a given biobank, we would fit a model:

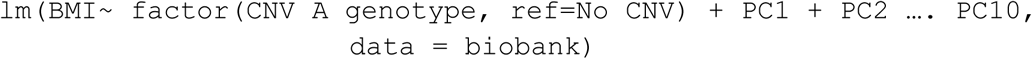

We then extracted the estimated regression coefficient, standard error, and p-value for each CNV. The regression coefficient represents the mean difference in sex and age corrected BMI or height, in standard deviations. Thus, if a CNV has an estimated main effect of 1.0 on BMI, carriers of that CNV have, on average, a BMI 1.0 standard deviations higher than those of the same age and sex. Finally, we performed a fixed effects meta-analysis using the R package metafor ^81^ to obtain an estimate of CNV effects across studies. We opted to use a fixed effects meta-analysis (as opposed to a random effects meta-analysis) because of the way in which the phenotypic measurements were scaled. Though there may be differences in demographics between the cohorts (for example, UKBB is older, MVP is male dominated), our scaling ensures that in each cohort, we are estimating the effect of any given variant *relative* to the rest of the cohort, which should be the same quantity across biobanks.

To calculate the variance explained by all recurrent CNVs, we fit a model containing all recurrent CNV loci coded as factors (+ covariates), and a covariate only model. We then meta-analysed R^2^ values with a 3-step process. We first applied Fisher’s transformation to the square root of R^2^ estimates. This stabilizes the variance and makes the sampling distribution approximately normal. We then applied the same fixed effects meta-analysis as above. Finally, the pooled estimate was transformed back into an R2 by reverting Fisher’s transformation and squaring the result. To obtain the variance explained by CNVs across studies, we compared the pooled R^2^ from the CNV + covariates model to the covariates only model, similar to^24^.

#### Psychiatric Medication Tabulation

The psychiatric medications, their RXNorm IDs, and the broad medication categories they belong to are summarized in Supplementary Table 8. A list of medications prescribed to each individual (lifetime use when available, self-reported use when not) was extracted within each dataset, limited to the medications listed in Supplementary Table 8. Individuals were placed into the “antidepressant” category if any of the medications labeled anti-depressant were included in their list. The same procedure was followed for antipsychotics and mood-stabilizers. Finally, the “all medication” category was generated taking the union of the three sub-categories (antidepressant, antipsychotic, and mood stabilizers).

#### Cross-Sectional Mediation Analysis

Mediation analysis was conducted with the `mediation`^82^ package in R, using raw phenotype values. The mediator model was:

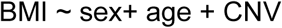

The outcome model was

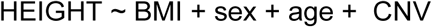

The relative contributions of direct and putative indirect effects on height were estimated via bootstrap with 1000 repetitions.

## Supplementary Information

### Extended Data

**Extended Data Figure 1:**
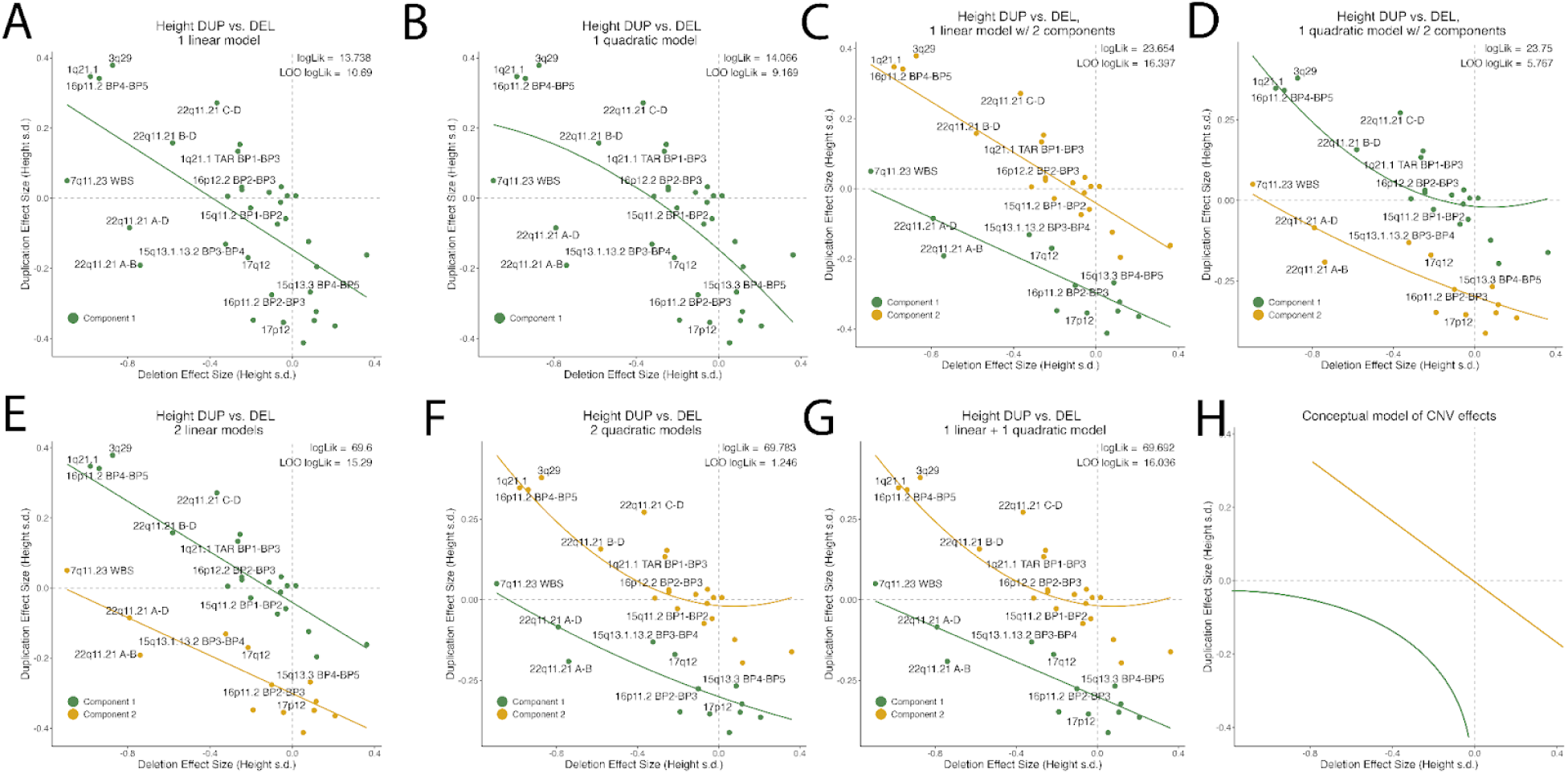
Effect sizes for reciprocal deletion and duplication on Height are consistent with two dose response curves. A model consisting of a **(A)** A linear model **(B)** a quadratic model **(C)** two linear models with shared parameters **(D)** two quadratic models with shared parameters **(E)** two linear models with distinct parameters **(F)** two quadratic models with distinct parameters **(G)** a linear and a quadratic model. LOO log-likelihood was calculated for each model fit to determine out of sample prediction accuracy for each model. Panels C, E, and G had the highest LOO log-Lik, suggesting two underlying dose-response relationships **(H)** We proposed a model in which the data consist of 1 linear component and 1 non-linear component.

**Extended Data Figure 2.**
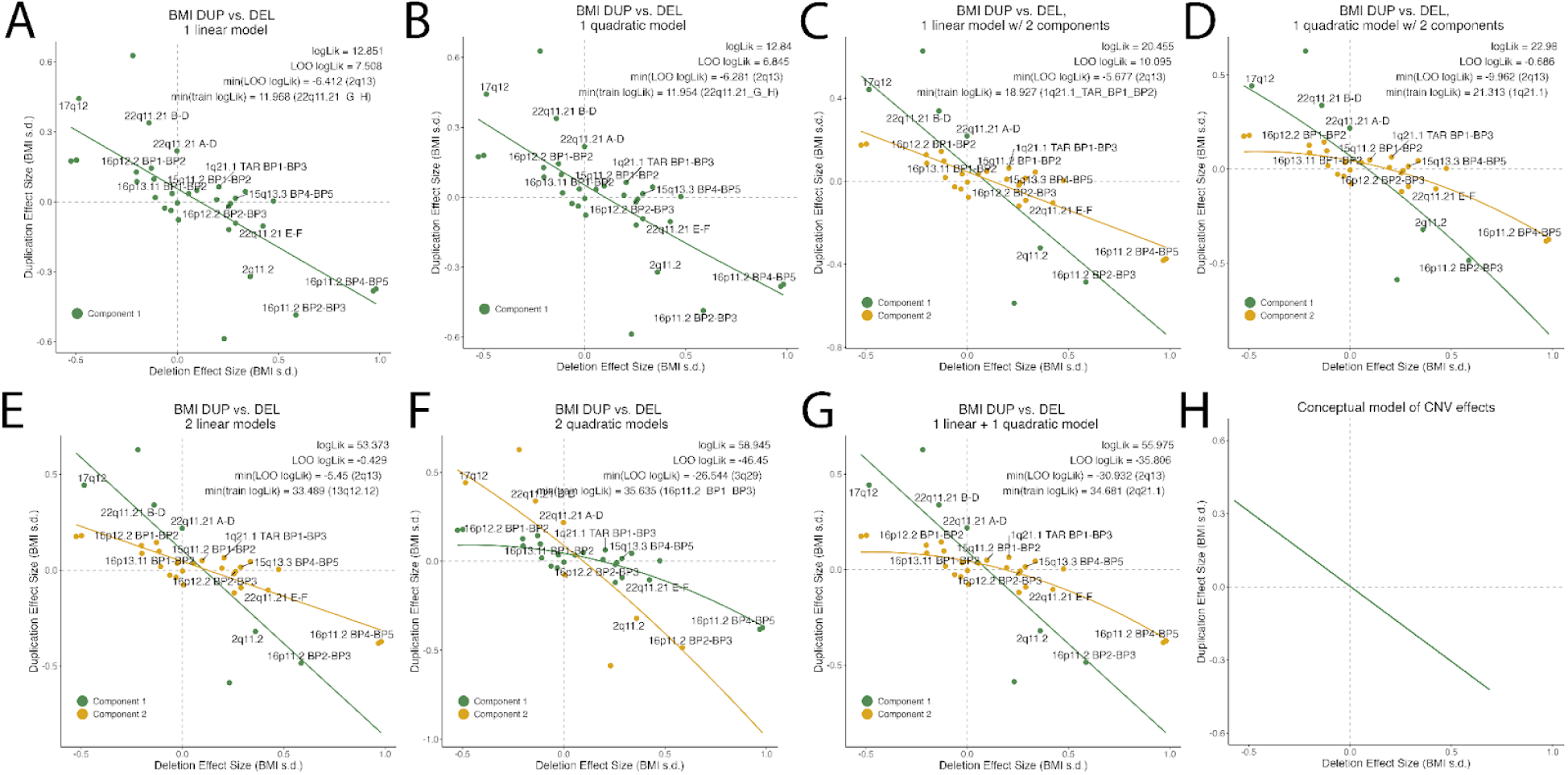
Effect sizes for reciprocal deletion and duplication on BMI are consistent with a single distinct dose response curve. A model consisting of a **(A)** A linear model **(B)** a quadratic model **(C)** two linear models with shared parameters **(D)** two quadratic models with shared parameters. BIC is comparable across A-D, suggesting that there is only one underlying model. **(E)** two linear models with distinct parameters **(F)** two quadratic models with distinct parameters **(G)** a linear and a quadratic model. **(H)** We proposed a model in which the data consist of 1 linear component.

**Extended Data Figure 3:**
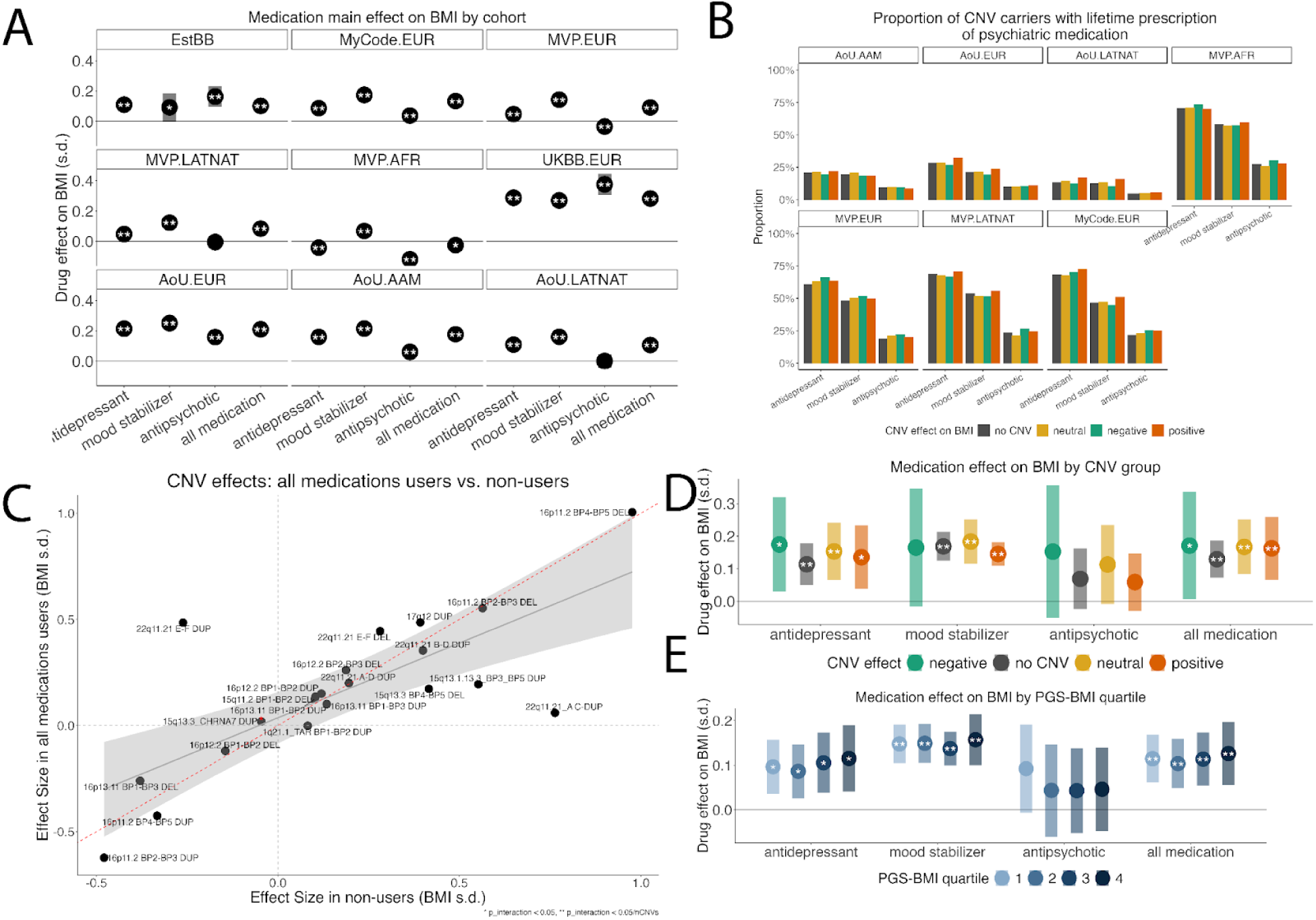
Additive effects of CNVs and psychiatric medication on body mass index. A). The effect of lifetime history of use of three broad classes of psychiatric medication (and their union) on BMI in all five biobanks. B). Proportion of individuals who reported lifetime use of three broad classes of medication, in CNV carriers and non-CNV carriers, in each biobank. C). Main effect of recurrent CNVs with and without controlling for medication use. D). The effect of three broad classes of psychiatric medication (and their union) on BMI by CNV background. White asterisks represent the effect of medication in that group (* = p < 0.05, ** = p < 0.001). Black asterisks represent an interaction between CNV genotype and medication on BMI. E). The effect of three broad classes of psychiatric medication (and their union) on BMI by PRS quartile. Asterisks are the same as in D. Summary statistics for panels A-E are in Supplementary Table 6

**Extended Data Fig. 4:**
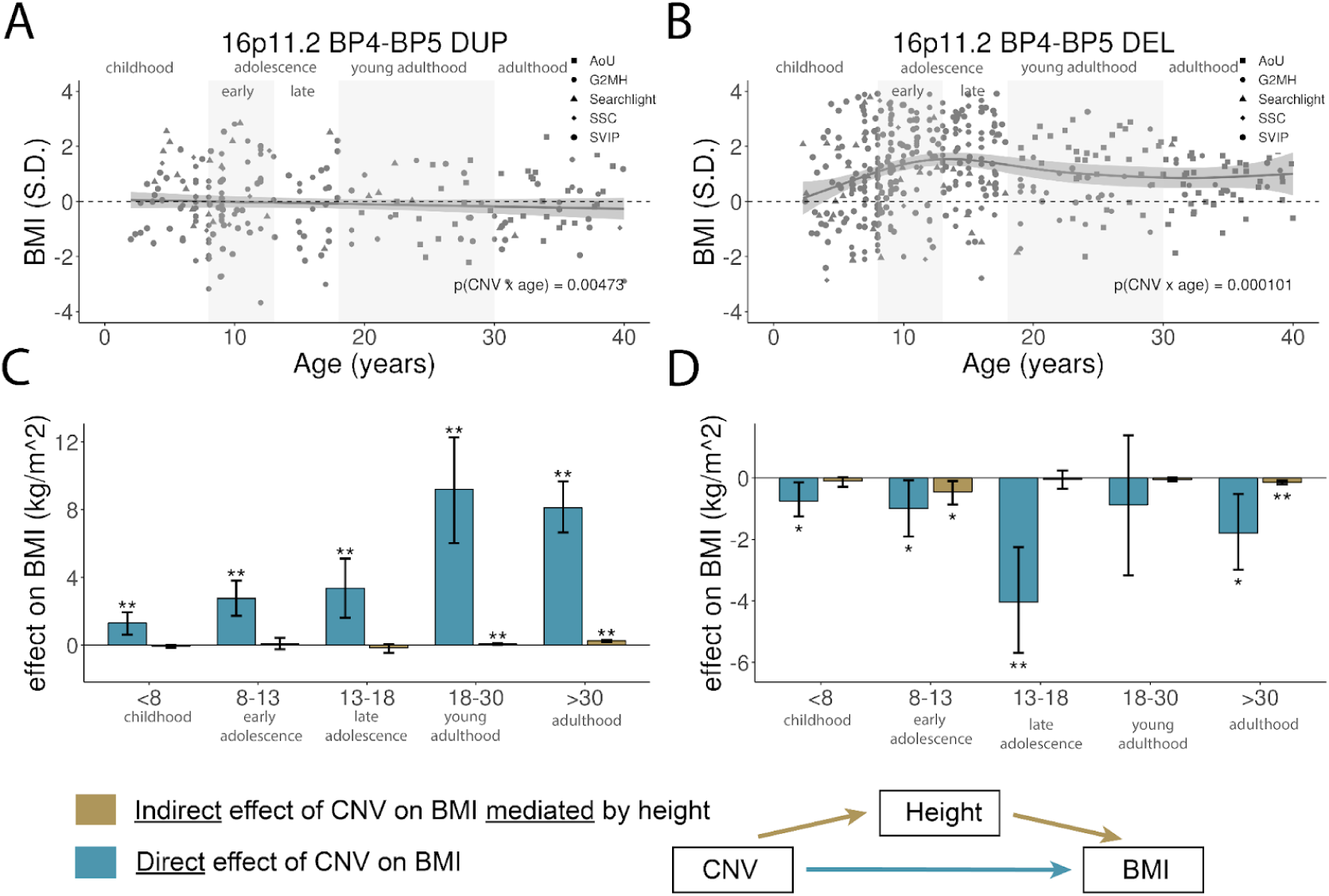
Effects of 16p11.2 BP4-BP5 CNVs on BMI differ by age. Age and sex normalized BMI values vs. age for (A) 16p11.2 BP4-BP5 DUP and (B) 16p11.2 BP4-BP5 DEL C-D). Age-stratified cross-sectional mediation analysis characterizing the distinct causal pathways underlying CNV effects on BMI, for DUP (C) and DEL

**Extended Data Fig. 5.**
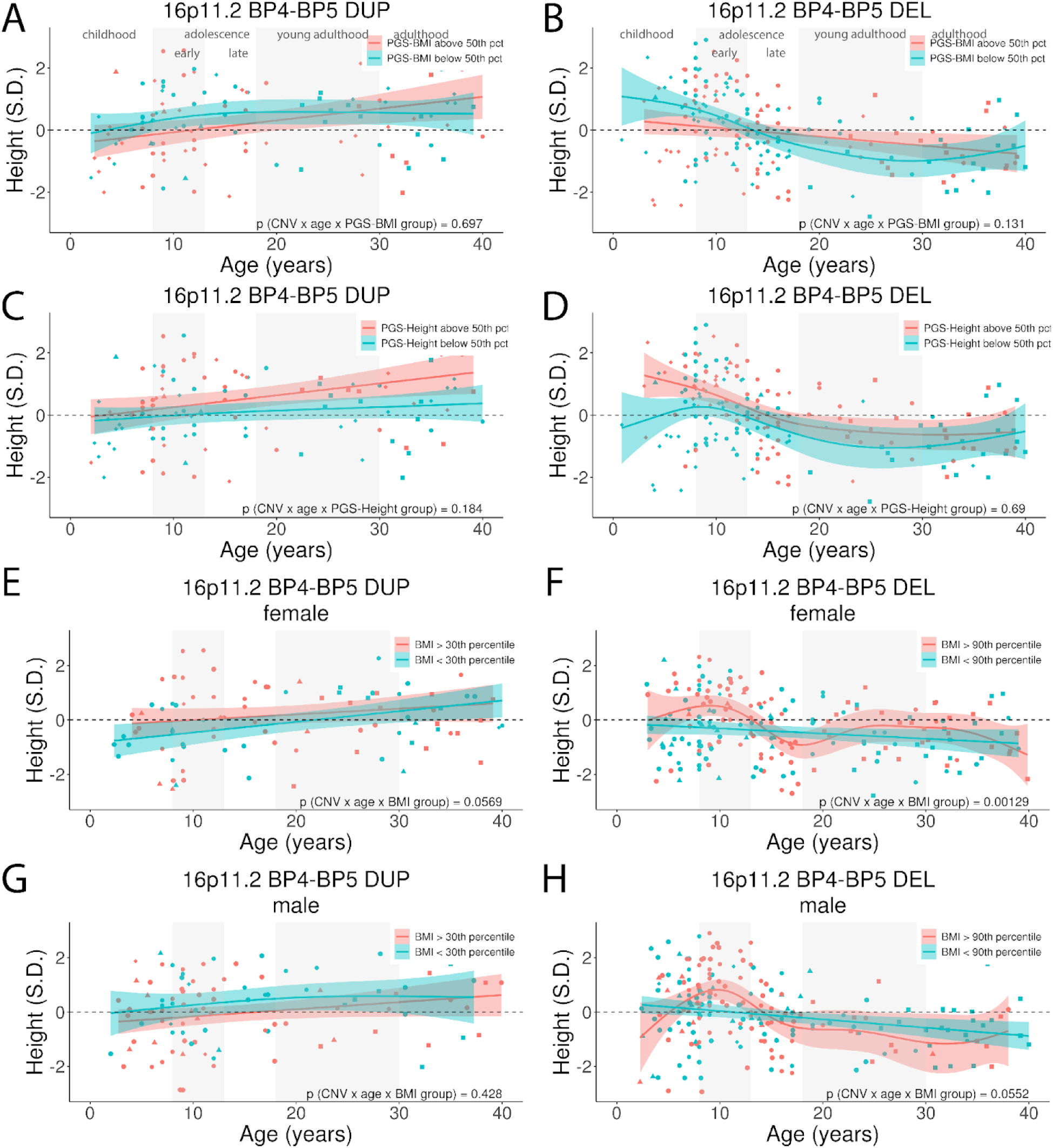
Age-dependent effects of 16p11.2 CNVs are not mediated by polygenic scores or sex. Trajectories stratified by (A-B) PGS-BMI (C-D) PGS-Height. (E-F) BMI (females only), and (G-H) BMI (males only). 16p11.2 BP4-BP5 DUP (A, C, E) and DEL (B, D, F).

**Extended Data Fig. 6:**
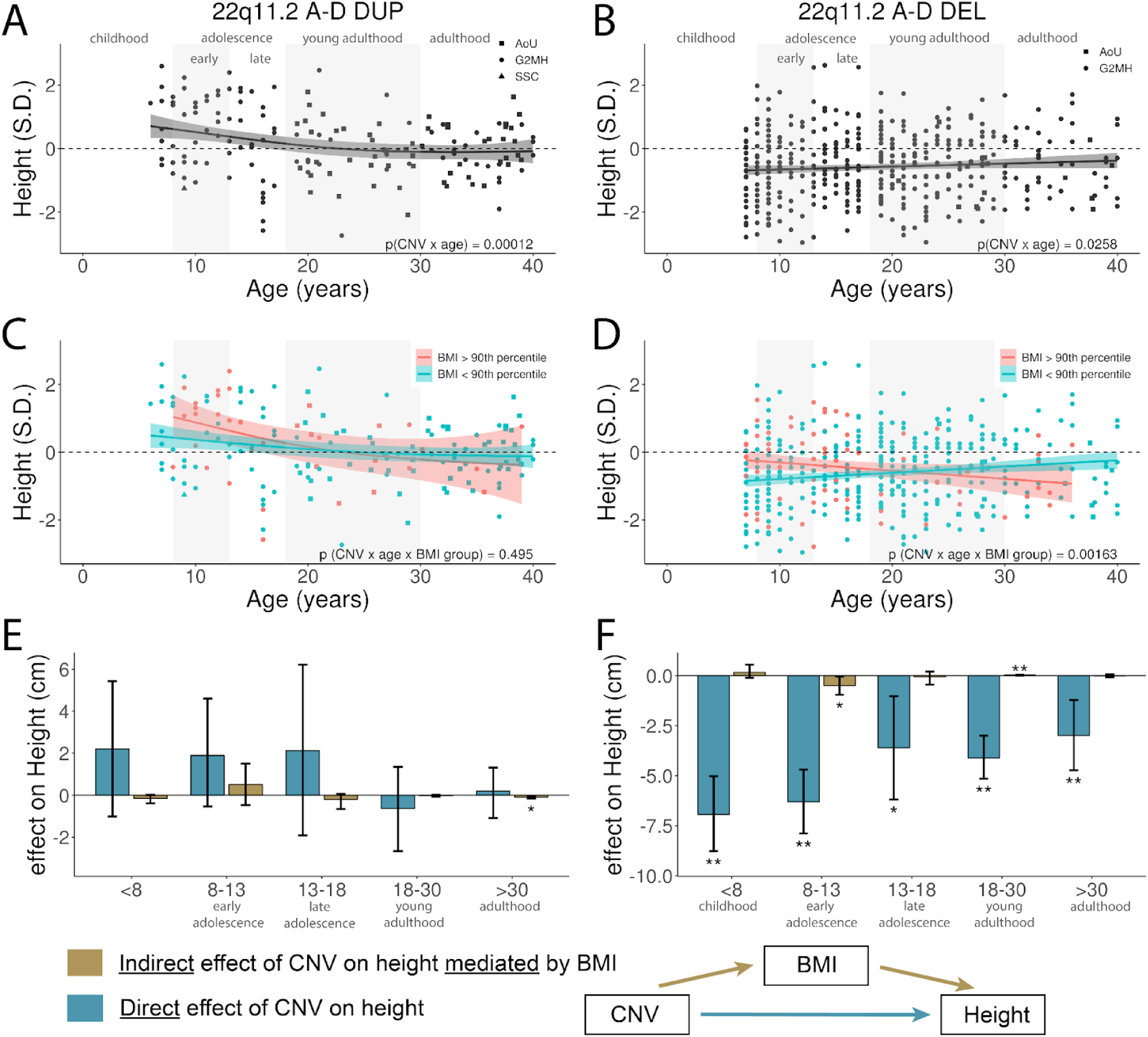
Effects of 22q11.21 A-D CNVs differ by age. Age and sex normalized height values vs, age for (A) 22q11.21 A-D DUP and (B) 22q11.21 A-D DEL. C-D). Cross sectional height trajectories further stratified by BMI at the 90th percentiles for DUP (C) and DEL (D). E-F). Age-stratified cross-sectional mediation analysis characterizing the distinct causal pathways underlying CNV effects on height, for DUP (E) and DEL (F).

**Extended Data Figure 7:**
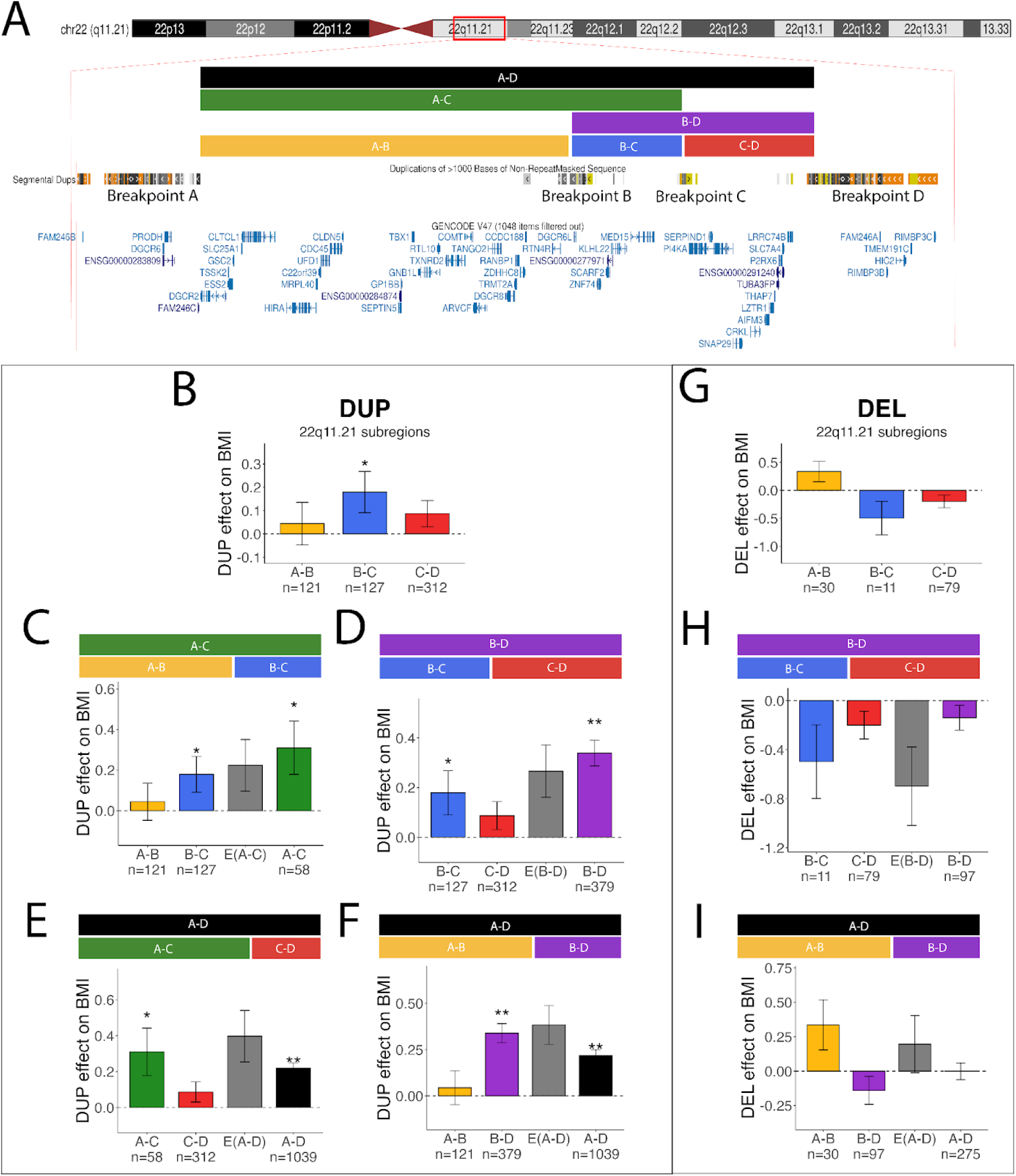
Dissection of the 22q11.2 A-D Locus and its effects on BMI. A). Schematic of the genes encompassed by the most commonly observed CNV breakpoints in our study, as defined by Gencode V49 accessed through the UCSC Genome Browser^56^ B-C). Breakdown of the effects of DUPs (B-F) and DELs (G-I) of 22q11.2 subregions. E(A-C) denotes the expected value of the effect size of A-C based on the sum of the effects of subregions A-B and B-C. Summary statistics for panels B-I are in Table S3.

**Extended Data Fig. 8:**
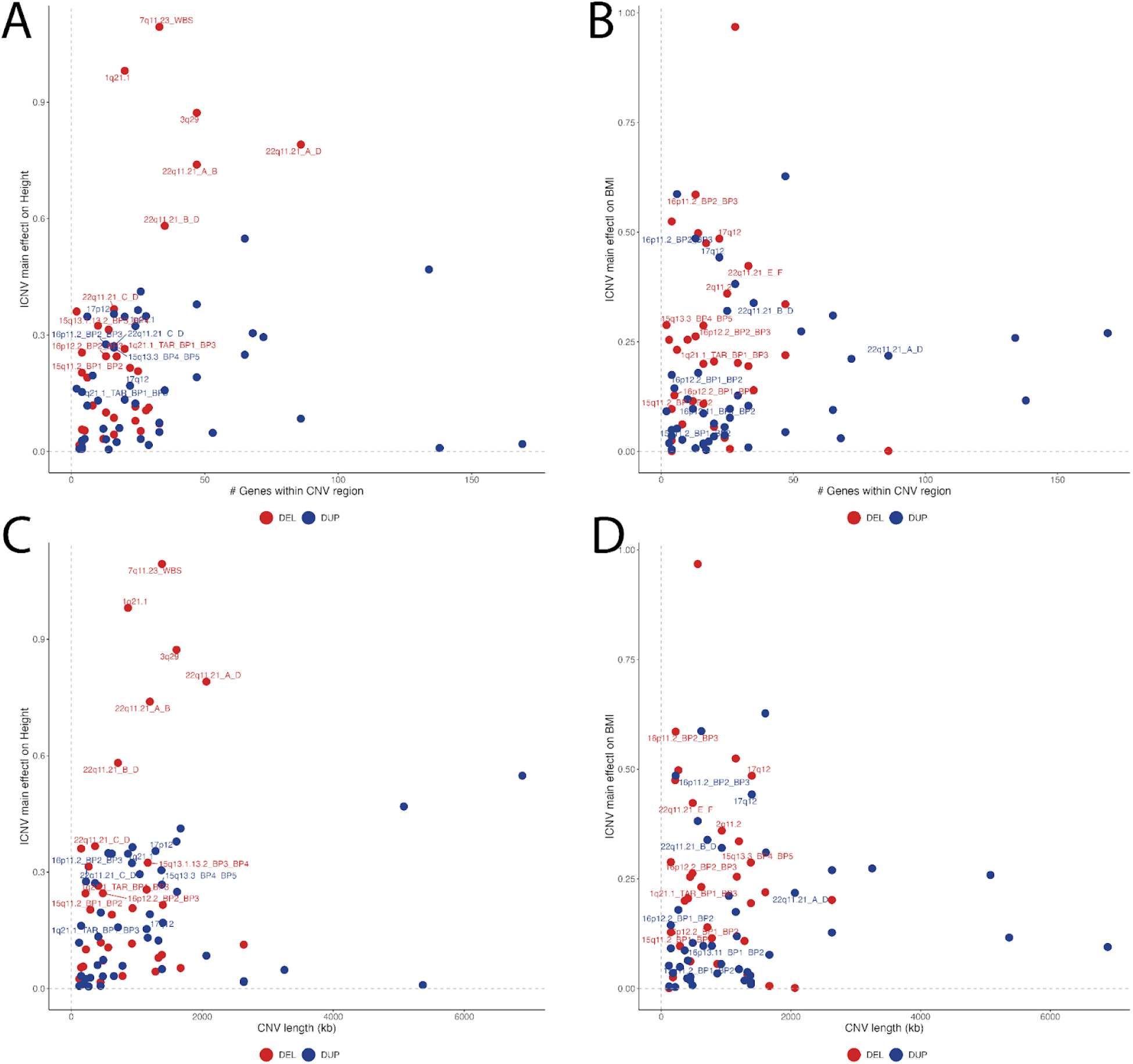
A-B). |CNV effect size| on (A) height and (B) BMI vs. number of genes within CNV region. CNVs with bonferroni significant main effects are labeled. C-D). |CNV effect size| on (C) height and (D) BMI vs. length of CNV region in base-pairs. CNVs with bonferroni significant main effects are labeled

### Supplementary Note 1: Model Misspecification

When quantifying genetic or environmental effects on a phenotype with multiple linear regression, there are a few assumptions that must be made in order to assess significance and create confidence intervals for effect size estimates. These assumptions are:

1. Linearity: the response (phenotype) *Y* is normally distributed about a *linear* combination of the predictor(s) *X*.
2. Homoscedasticity: the variance of the residuals are equal across *X*
3. Normality of residuals: the residuals are normally distributed
4. Independence of errors: the *y*_i_ are independent from each other… i.e., the phenotype of one sample does not influence the phenotype of another sample
5. Absence of multicollinearity: the predictors *X* are uncorrelated with each other

When these assumptions are broken, we say that the model is “misspecified”, which means that the estimators of the model parameters and their corresponding test statistics do not behave as we would expect them to, which may lead us to incorrectly accept or reject a null hypothesis about the effect of a predictor variable. The effect of model misspecification can vary between parameters and their estimators, and also depends on which assumptions were broken and how they were broken.

There are a few characteristics of genetic and phenotypic data that make genetic studies particularly vulnerable to model misspecification. First, phenotypic data is rarely perfectly normally distributed, which can lead to conflicts with assumptions 2 and 3. ^20^ discuss the limits of the normal approximation for height, and demonstrate that a log-normal approximation may be more accurate. Second, when studying individuals in a population, it is rare that errors are independent (assumption 4), especially when closely related individuals are involved in the same study. Beyond genetic relationships between individuals, genetic variants are often correlated *within* an individual due to either linkage disequilibrium (LD), population stratification, or assortative mating, which conflicts with assumption 5.

With real world data, it is unlikely that a linear model will be perfectly specified, and it is usually sufficient for the assumptions to be approximately satisfied. However, there is evidence that the effects of model misspecification can be amplified when estimating interaction terms, which will be discussed at length in the following sections. In this work, we are particularly interested in estimating non-additive (or interaction) effects between genetic factors. Thus, it is imperative that we understand the effects of model misspecification on the interaction estimates in this study, and apply the appropriate methods to avoid misspecifying the model in the first place.

#### Non-normality of the phenotype of interest

A common theme that has arisen in previous work investigating non-additive genetic effects on BMI and height ^5,20,83^ is that applying the normal approximation to non-normal phenotypes can result in type 1 errors. Non-normality can often be resolved by applying one of a few transformations to the data. A common transformation is the non-parametric rank based inverse normal transformation (RINT), which ranks the phenotype values and fits their quantiles to a normal distribution. ^84^ demonstrated that applying this transformation ameliorates the non-normality issue when testing for interactions of common SNPs and age on BMI. However, ^85^ demonstrated that applying an INT can reduce power to detect epistatic interactions. Due to the already limited power in our study (see Supp Note 2), we decided that a parametric transformation would be more appropriate. In their study of genetic influence within-person longitudinal change in anthropometric traits in UKBB, ^5^ applied a BoxCox transformation to remove the strong mean-variance relationship in BMI and other traits. We chose this transformation for our study and applied it as in ^5^ (see methods). All results discussed in the main text use this transformation.

In the context of interactions between rare variants of large effects and PGS, even small deviations from the normal approximation can lead to *biased* estimates of interaction terms, which can lead us to make conclusions about these interactions that are entirely scale dependent. By the central limit theorem, a PGS is normally distributed, as it is the sum of many independent effects. If the phenotype of interest is not normally distributed, a normally distributed PGS *cannot* have a linear relationship with the phenotype while maintaining homoscedasticity, as there is no linear transformation of a normal random variable that is non-normal. This remains true when we introduce a variant of large effect (CNV) to the model… the distribution of the effect of CNV + PGS remains normal when we add the constant CNV effect. However, introducing an interaction term allows the PGS effect to vary depending on CNV background. If the relationship between PGS and the phenotype is non linear, then the variance in the phenotype that can be explained by the PGS will vary across the phenotype distribution. As the magnitude of the CNV effect increases, the effect of the PGS will change with respect to its effect in someone with no CNV, since the variance in phenotype is not constant. Thus, CNVs with large effects will appear to have large interaction effects with the PGS.

This all sounds very theoretical, but if we aren’t careful, it can lead us to some very false conclusions, especially in the rare variant space. To illustrate this, we fit two CNVxPGS interaction models for 29 well-powered CNVs in the UKBB for both height and BMI. For height, we used the 1) normally distributed BoxCox transformed phenotype values (which are very similar to the raw height values), and 2) an additional set of phenotype values obtained by squaring the BoxCox values (and keeping the sign). This height distribution has much heavier tails than the normal distribution. For BMI, we used 1) the normally distributed BoxCox transformed BMI values, and 2) the raw BMI values, which have a strong rightward skew. We observed that the main effects of the CNVs were not very sensitive to these transformations. However, the change in interaction effect estimates increased as the magnitude of the CNV main effect increased. For height, the estimates were biased positively, as there is more variance in the tails of the squared height distribution than the normal distribution. This could lead one to conclude that “PGS tends to have a stronger effect in those carrying CNVs with large effects on height”. For BMI, the estimates were biased negatively for CNVs that decreased BMI, since there is less variance in the lower end of the raw distribution compared to the normal distribution. The estimates were biased positively for CNVs that increased BMI, since there is more variance in the upper end of the raw distribution than the normal distribution. This may lead one to conclude that the interactions are “synergistic”, where the strength of the PGS as a predictor of BMI changes linearly as a function of rare variant background. These results demonstrate that in the rare variant space, model misspecification resulting from phenotypes with non-normal distribution can lead to conclusions about the nature of genetic interactions that are scale dependent, and that this effect increases as rare variant effect increases.

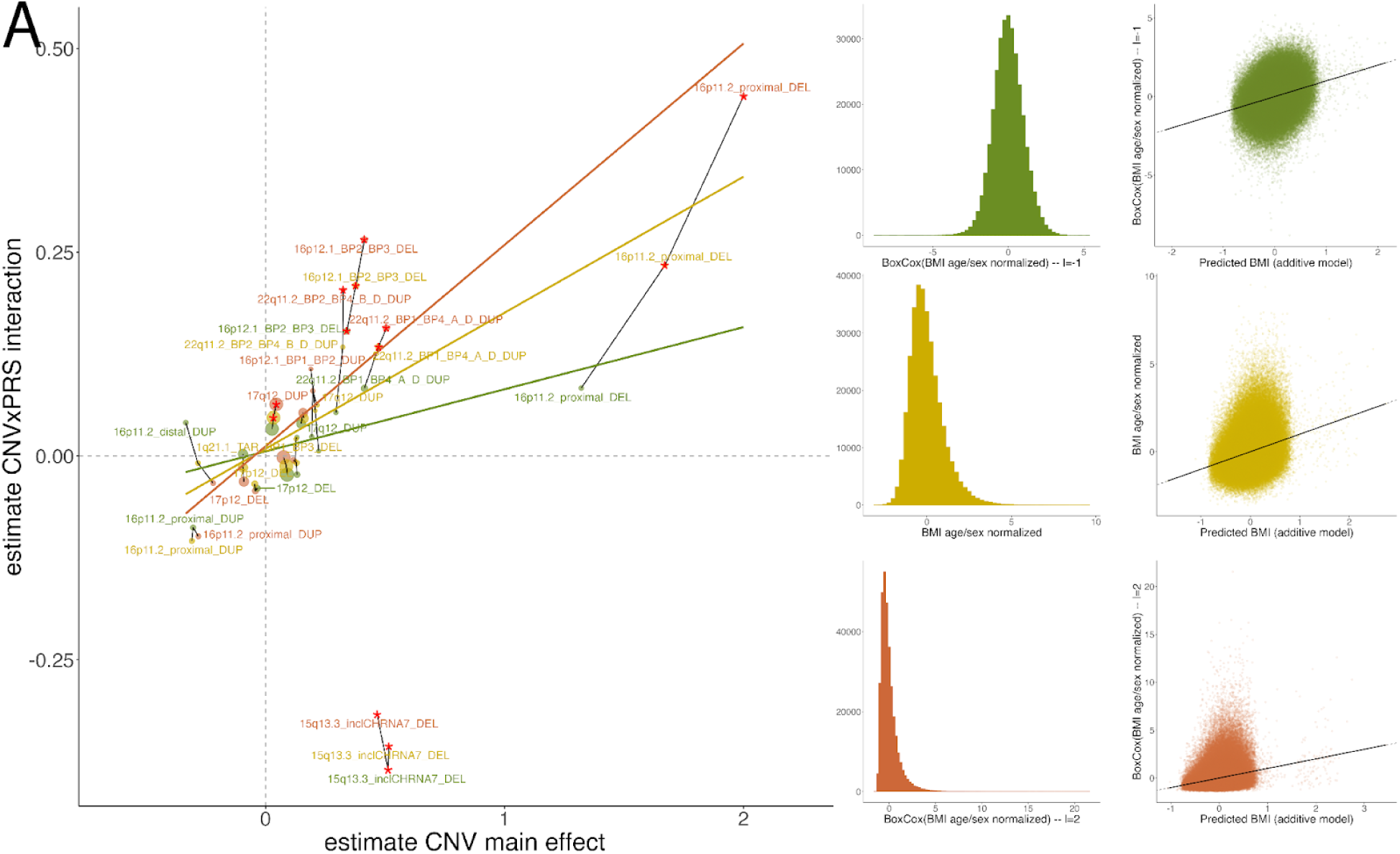

#### Linkage disequilibrium, population stratification, and assortative mating

A second manner in which genetic studies are especially prone to model misspecification is that genetic variants are often correlated with each other, whether it be on the same chromosome through LD, or across chromosomes through population stratification and assortative mating. It is widely accepted that SNP effect size estimates in a GWAS are inflated due to LD, and this is often a consideration when constructing PGSs from GWAS results (addressed by LD pruning). There is not a clear consensus on how to handle LD in studies of statistical epistasis in the SNP space. One paper conducted a genome wide scan for epistasis between SNPs on a variety of traits, and reported that most of their significant hits were between pairs of SNPs within 1Mb of each other ^86^. Other studies removed SNP pairs that are known to be in LD^87^. For the rare variants in our study, which likely arise de Novo, LD with SNPs is not a concern, though they may still be subject to the effects of population stratification or assortative mating if they were inherited. It has been demonstrated that cross chromosome correlations between SNPs (population stratification or assortative mating) can artificially inflate marker-based heritability estimators ^88^, though there are no studies to our knowledge that address this issue in the context of detecting epistasis. A common practice is to “control” for population stratification by including ancestry principal components when regressing phenotype on genotype (as we did in this study, see methods), but this does not address the effects of population stratification on estimates of interactions between correlated genetic factors.

### Supplementary Note 2: Statistical Power

Even if our model is perfectly specified (see Supp Note 1), there is still the issue of statistical power. Power to detect both main effects and interaction effects was estimated using a Monte Carlo simulation based method. A population of 1 million with *n* CNV carriers (categorical) were generated for each simulation. PGSs and sex were randomly assigned (assuming no assortative mating). Next, a linear model was used to generate phenotypic values based on the specified CNV main effect or interaction term, and normally distributed residual variance depending on the inclusion of PGS in the model (which was assumed to have R^2^=0.1), for a total variance of 1 and mean of 0. Finally, main effect or interaction terms were estimated with linear regression. This was repeated 1000 times for each set of parameters, and power was calculated as the proportion of tests that detected the true underlying effect with alpha=0.05.

We are well powered to detect CNV main effects of +/- 0.85 S.D. with 10 CNV carriers. The majority of main effect point estimates in our study were within this range, with a few CNVs having main effects closer to 1 S.D. Thus, we included all CNVs in our main effect estimates that had at least 10 carriers in at least one cohort. Estimates of CNV main effects generated from fewer than 10 carriers in a single cohort were unstable, so those were excluded.

CNV-PGS interactions are expected to be of a much smaller magnitude than main effects. An interaction effect that exceeds the main effect of the PGS (0.297 S.D. for BMI, 0.597 S.D. for height) is unlikely, since that would mean the PGS effect is either doubled or completely negated in CNV carriers. Since the expected interaction term is lower, our power was more limited in this context. Based on our power calculations below, we only included CNVs with over 200 carriers in all cohorts combined in our interaction tests.

For a categorical X categorical interaction where one category is quite common (i.e., sex or medication use), power was slightly greater than categorical X continuous, since we only need to detect a difference in CNV effect between two equally sized groups. However, it is unlikely again that this interaction is larger than the CNV main effect itself (ie, CNV effect is doubled or completely negated in females vs. males). Thus, we elected to use the same 200 carrier cutoff in testing CNVxSex interactions.

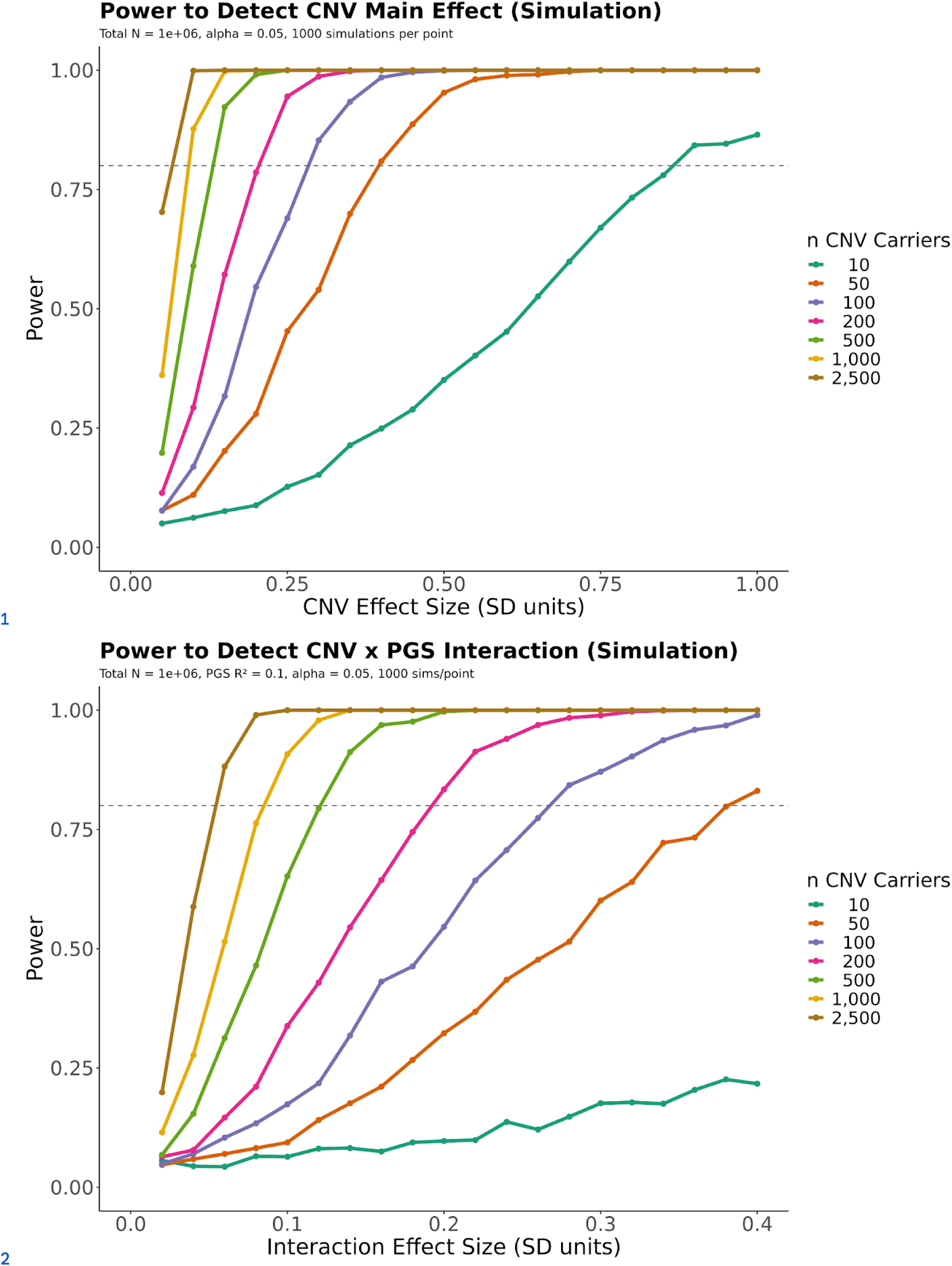

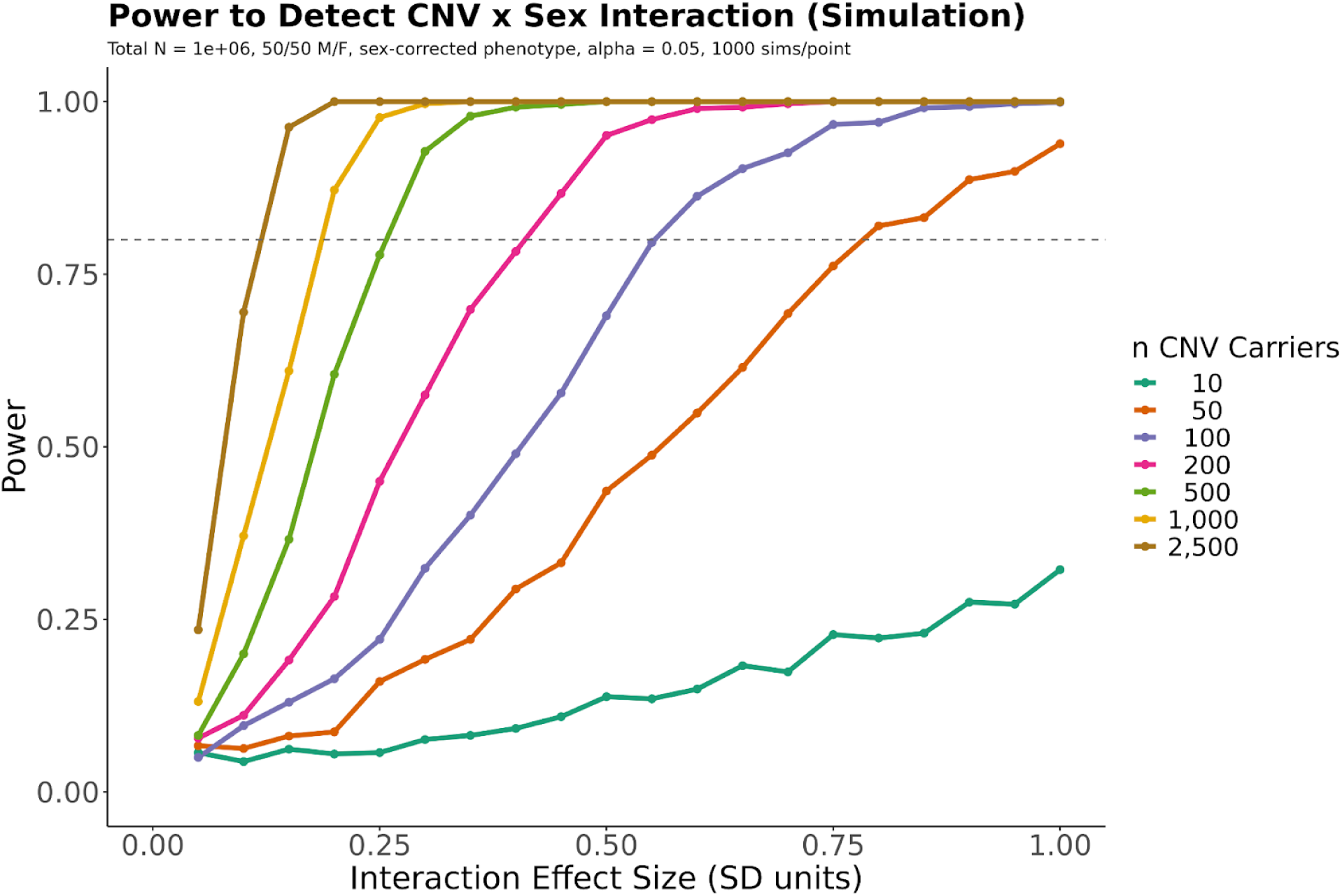

### Supplementary Note 3: Comparison of our results with Milind et al

Theoretical model of dose-response curves by Milind et al.^58^ emphasizes that non-monotonic gene-dosage response curves may be a common property of height-associated loci, such that both deletions and duplications typically shift the trait in the same direction (**Millind et al Fig 1F**). Our results show that a subset of loci for height (∼30%) exhibit asymmetric effects, and subset of these (e.g. 22q11.2 A-B and 22q11.2 A-D) are non-monotonic such that effects are negative for both DEL and DUP. However, our data suggest that a U-shaped non-monotonic relationship is not an intrinsic property of most loci. Some examples of buffering appear to be explainable by the aggregate behavior of multiple genes within the A-D a locus. Attenuation of the A–D DUP effect is consistent with partial (additive) cancellation of opposing effects from A–B and B–D subregions (**Fig. 6F**). Conversely, when the combined effect of A-B and B-D DELs, both of which are in the negative direction, we see a sub-additive effect (**Fig. 6I**). Thus effects appear to be attenuated in the positive direction for DUP and attenuated in the negative direction for DEL. To summarize, our results are consistent with results from Milind et al in some respects: gene dosage effects on height show evidence of buffering, and effects are biased in the negative direction. Our results contrast in other respects: Non-monotonicity is not a general property of the trait, it is restricted to a specific subset of loci. Buffering appears to involve combinations of multiple genes, and we see evidence that buffering can stabilize effects in both directions.

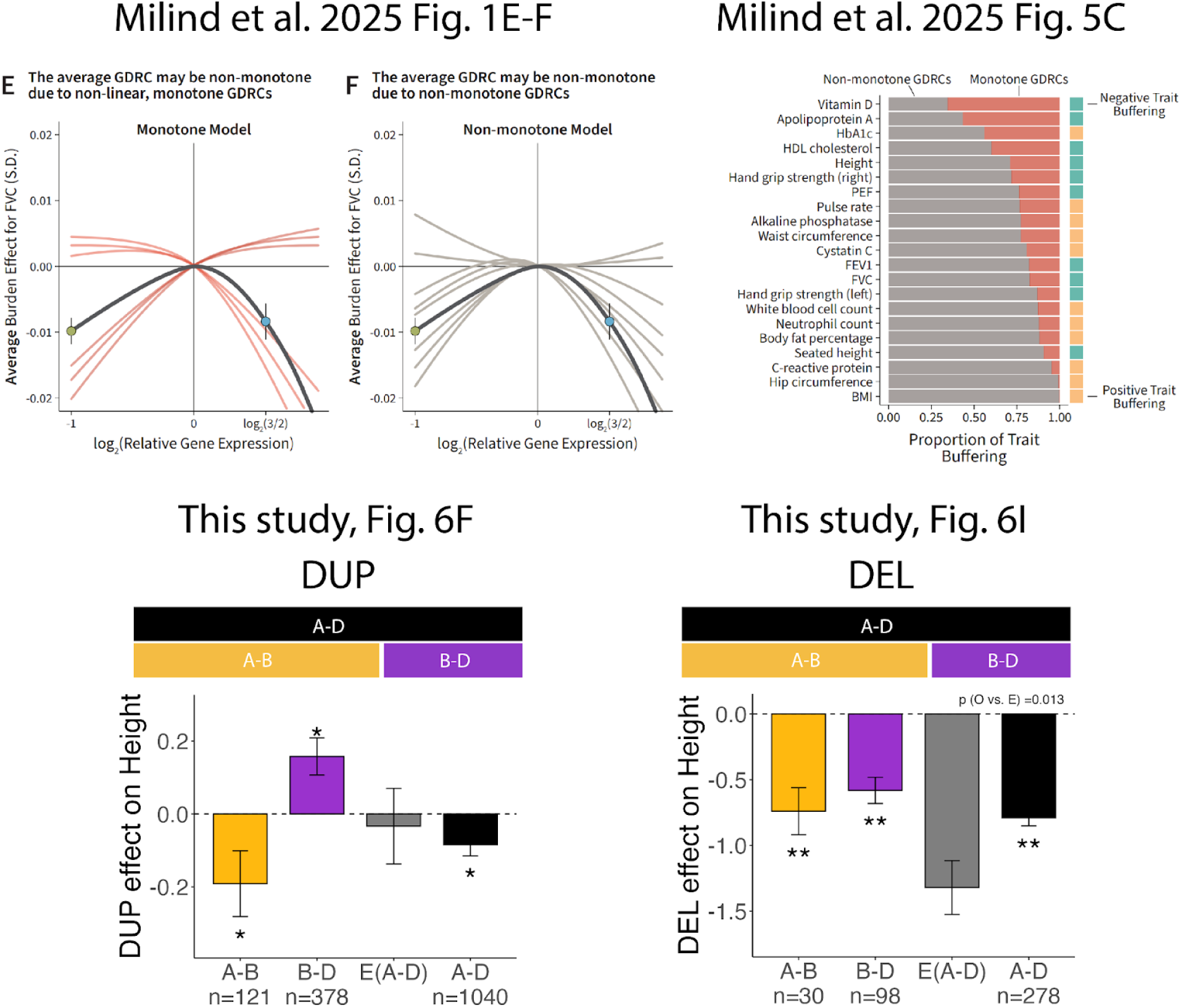

## Code Availability

Code for shared analysis pipeline and meta-analysis can be accessed on GitHub: https://github.com/mollysacks/Sacks2026

## Notes

### Competing Interest Statement

The authors have declared no competing interest.

### Author Declarations

IRB of University of California San Diego gave ethical approval for this work

